# Parkinson’s Disease Pathogenic Variants: Cross-Ancestry Analysis and Microarray Data Validation

**DOI:** 10.1101/2024.12.16.24319097

**Authors:** Samantha Hong, Mathew J. Koretsky, Jens Lichtenberg, Hampton Leonard, Vanessa Pitz, the Global Parkinson’s Genetics Program (GP2)

## Abstract

**Background:** Known pathogenic variants in Parkinson’s disease (PD) contribute to disease development but have yet to be fully explored by arrays at scale.

**Objectives:** This study evaluated genotyping success of the NeuroBooster array (NBA) and determined the frequencies of pathogenic variants across ancestries.

**Method:** We analyzed the presence and allele frequency of 34 pathogenic variants in 28,710 PD cases, 9,614 other neurodegenerative disorder cases, and 15,821 controls across 11 ancestries within the Global Parkinson’s Genetics Program dataset. Of these, 25 were genotyped on NBA and cluster plots were used to assess their quality.

**Results:** Genes previously predicted to have high or very high confidence of causing PD tend to have more pathogenic variants and are present across ancestry groups. Twenty-five of the 34 pathogenic variants were typed by the NBA array and classified “good” (n=12), “medium” (n=4), and “bad” (n=9) variants.

**Conclusion:** Our results confirm the likelihood that established PD genes are pathogenic and highlight the importance of ancestrally diverse research in PD. We also show the usefulness of the NBA as a reliable tool for genotyping of rare variants for PD.

## Introduction

With the expanding global burden of Parkinson’s disease (PD), investigating causes and risk factors becomes increasingly important. Most cases of PD occur sporadically and are thought to arise from a complex interplay of genetic and environmental factors (1–3). Pathogenic variants can be common or rare, and often cause loss or gain of function in critical proteins, increasing the risk of developing disease. Identifying these genetic contributors can provide insights into disease mechanisms and potential therapeutic targets. In PD, pathogenic variants have been found in well-known genes such as *LRRK2, PRKN, PINK1*, and *SNCA*, which have been linked to both familial and sporadic forms of the disease (4–6).

Rare disease-causing variants have been identified through linkage studies of familial PD (7). Genotyping technologies allowed for the determination of the genetic makeup of individuals, and recent developments in next-generation sequencing (NGS) techniques, such as exome and genome sequencing, have facilitated the discovery of pathogenic variants. Through a recent PD genome-wide association study (GWAS) meta-analysis, 90 independent risk variants were uncovered in individuals of European ancestry (8). Though there have been recent efforts to conduct genetic studies for non-European populations, such as in East Asian (9), Latino (10), and African (11) ancestries, it is imperative to continue expanding our knowledge of the genetic influence of PD in underrepresented ancestries for a better understanding of the disease as a whole.

A review by Blauwendraat et al. categorized 21 genes with reported pathogenic variants according to their likelihood of causing PD (12). Research in PD genetics has been primarily focused on samples of European ancestry (13), with Blauwendraat et al. results reflecting this lack of diversity. The NeuroBooster array (NBA) is a custom designed platform aimed to screen for variants of neurological conditions across diverse populations (14). We aim to evaluate genotyping success of the NBA (14) and analyze frequencies of pathogenic variants in diverse populations leveraging data from the Global Parkinson’s Genetics Program (GP2).

## Methods

### The Global Parkinson’s Disease Genetics Program

Data used for this study were obtained from GP2. GP2 aims to improve understanding of the genetic architecture of PD across globally diverse populations. This study utilized samples in GP2 release 7 to investigate pathogenic variants that have been studied in a limited number of ancestral groups across a more diverse set of populations. The data consisted of 28,710 PD cases, 9,614 other neurodegenerative disorder (NDD) cases, and 15,821 controls across 11 ancestry groups: African American (AAC) (2.05%), African (AFR) (4.88%), Ashkenazi Jewish (AJ) (4.90%), Admixed American/Latin American (AMR) (1.19%), Complex Admixture History (CAH) (1.57%), Central Asian (CAS) (1.67%), East Asian (EAS) (9.54%), European (EUR) (71.73%), Finnish (FIN) (0.21%), Middle Eastern (MDE) (1.07%), and South Asian (SAS) (1.17%) (15). More demographic information is included in Supplementary Table 1.

### Data processing

Our study looked at variants in the 21 PD genes included in the Blauwendraat et al. analysis: *SNCA, PRKN, UCHL1, PARK7, LRRK2, PINK1, POLG, HTRA2, ATP13A2, FBX07, GIGYF2, GBA1, PLA2G6, EIF4G1, VPS35, DNAJC6, SYNJ1, DNAJC13, TMEM230, VPS13C, and LRP10*. All variants in the GP2 imputed data (imputation R^2^ threshold of 0.3) were annotated across all ancestries using ANNOVAR (version 2020-06-07) and the ClinVar database (version 2024-09-17). Variants labeled as “Pathogenic” in the “Clinical Significance” field within the genes of interest were extracted using PLINK 1.9 (16,17); the ClinVar ANNOVAR annotations for the 34 “Pathogenic” variants are included in Supplementary Table 2. PLINK was also used to calculate the frequencies of the pathogenic variants across ancestries. Data cleaning and wrangling was conducted using Python (3.10.12). We then produced cluster plots to assess genotyping quality of NBA on the GP2 release 7 samples. Cluster plots visualize the different allelic statuses (homozygous reference, heterozygous, homozygous alternate) by plotting Theta and R values that are provided by Illumina at the time of genotyping to visually assess the accuracy of the genotyping results (18,19). We used two criteria to assess genotype success of a variant based on clustering quality: 1) shape and distinction of the clusters and 2) number of samples for which the variant could not be called (NCs). A plot was labeled as “bad” if it appeared to not follow the normal “spread” in regards to the distance between clusters (change in Theta between clusters <= 0) and if a high number of NCs/low call rate was observed (NCs >= 10). If a plot exhibited expected behavior, it was classified as “good”. Plots that were difficult to classify or where analysts’ responses differed were labeled as “medium”. An overview of our workflow is shown in Figure 1.

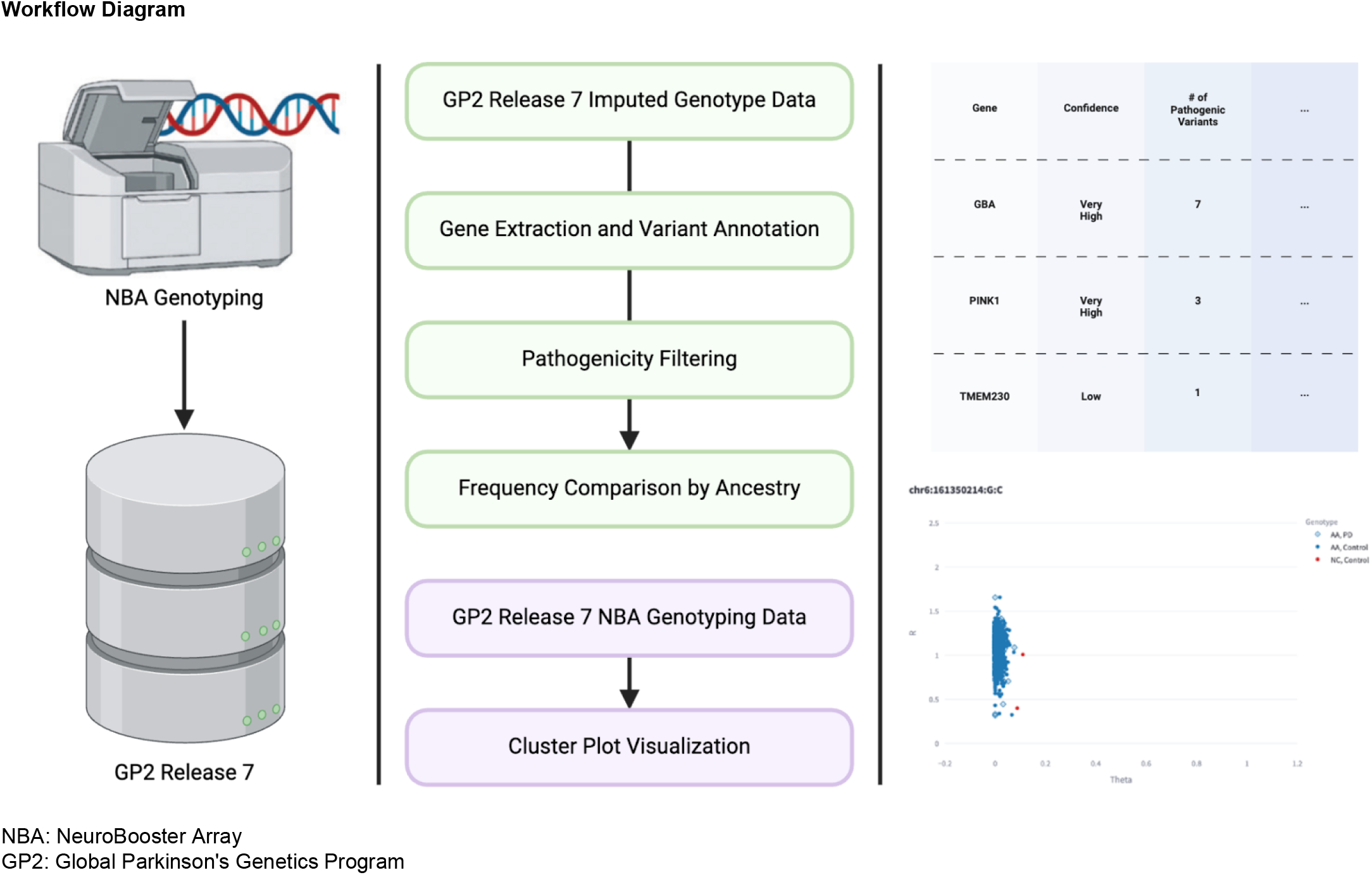

## Results

A total of 34 labeled pathogenic variants from the 21 genes of interest were analyzed (Supplementary Table 2). It must be noted that only 10 of the pathogenic variants within the established PD genes had a disease name that included some form of PD (denoted within the “CLNDN” field in ClinVar); the other 24 pathogenic variants had other disease names, such as Gaucher disease and Progressive Sclerosing Poliodystrophy, or was “not_provided”.

To assess the quality of genotyping pathogenic variants with the NBA, we created cluster plots for each non-imputed variant and had two analysts visually inspect them. Twenty-five of the 34 pathogenic variants were typed (non-imputed) and thus visualized through a cluster plot (as described in Methods). On average, variants with a lower MAF and missingness rate in GP2 release 7 had higher genotyping quality (Supplementary Figure 1).

After classification, there were 12 variants labeled as “good”, 4 labeled as “medium”, and 9 labeled as “bad”, as shown in Supplementary Table 3. The number of NCs for each typed variant is also included, along with the minor allele frequency and missiness rate for that variant in GP2 release 7. Images of cluster plots for all 25 typed variants can be found at https://github.com/GP2code/pathogenic_pd_variants_clusters (https://zenodo.org/records/14193209).

The distribution of the 34 pathogenic variants across the genes are listed in Table 1. Interestingly, there are four genes that were classified as having a “very high” confidence for causing PD by Blauwendraat et al., *FBXO7, SNCA, PARK7*, and *VPS35*, that had zero pathogenic variants found. Five of the six genes that were classified as “low” confidence for causing PD also had zero pathogenic variants present across all ancestries; *TMEM230* was the only “low” probability gene that had a pathogenic variant present. To further assess how well NBA captures pathogenic variants in these genes, we compared the pathogenic variants we found within each of these genes to the reported pathogenic variants in gnomAD (v4.1.0) (Supplementary Table 4). However, it must be noted that the “Pathogenic” variants within each gene from gnomAD may not be associated with a clinical diagnosis involving PD, per ClinVar.

Number of variants based on confidence in gene and ancestry.

| Gene | Confidence as<br>actual PD gene<br>per<br>Blauwendraat<br>et al. 2020 | Total number of<br>pathogenic<br>variants | AAC | AFR | AJ | AMR | CAH | CAS | EAS | EUR | FIN | MDE | SAS |
| --- | --- | --- | --- | --- | --- | --- | --- | --- | --- | --- | --- | --- | --- |
| ATP13A2 | very high | 5 | 0 | 1 | 0 | 1 | 0 | 0 | 1 | 3 | 0 | 0 | 0 |
| FBXO7 | very high | 0 | 0 | 0 | 0 | 0 | 0 | 0 | 0 | 0 | 0 | 0 | 0 |
| GBA | very high | 7 | 0 | 2 | 0 | 0 | 1 | 0 | 0 | 4 | 0 | 1 | 0 |
| SNCA | very high | 0 | 0 | 0 | 0 | 0 | 0 | 0 | 0 | 0 | 0 | 0 | 0 |
| PLA2G6 | very high | 3 | 0 | 0 | 0 | 1 | 0 | 1 | 1 | 1 | 0 | 0 | 0 |
| PINK1 | very high | 3 | 0 | 0 | 0 | 0 | 0 | 0 | 2 | 2 | 1 | 1 | 0 |
| PARK7 | very high | 0 | 0 | 0 | 0 | 0 | 0 | 0 | 0 | 0 | 0 | 0 | 0 |
| LRRK2 | very high | 1 | 0 | 0 | 0 | 0 | 0 | 1 | 0 | 1 | 0 | 1 | 0 |
| PRKN | very high | 5 | 0 | 0 | 1 | 1 | 1 | 0 | 1 | 1 | 0 | 0 | 0 |
| VPS35 | very high | 0 | 0 | 0 | 0 | 0 | 0 | 0 | 0 | 0 | 0 | 0 | 0 |
| POLG | high | 8 | 2 | 4 | 1 | 0 | 0 | 0 | 0 | 6 | 0 | 0 | 0 |
| DNAJC6 | high | 0 | 0 | 0 | 0 | 0 | 0 | 0 | 0 | 0 | 0 | 0 | 0 |
| SYNJ1 | high | 1 | 0 | 1 | 0 | 0 | 0 | 0 | 0 | 0 | 0 | 0 | 0 |
| VPS13C | high | 0 | 0 | 0 | 0 | 0 | 0 | 0 | 0 | 0 | 0 | 0 | 0 |
| HTRA2 | low | 0 | 0 | 0 | 0 | 0 | 0 | 0 | 0 | 0 | 0 | 0 | 0 |
| EIF4G1 | low | 0 | 0 | 0 | 0 | 0 | 0 | 0 | 0 | 0 | 0 | 0 | 0 |
| LRP10 | low | 0 | 0 | 0 | 0 | 0 | 0 | 0 | 0 | 0 | 0 | 0 | 0 |
| DNAJC13 | low | 0 | 0 | 0 | 0 | 0 | 0 | 0 | 0 | 0 | 0 | 0 | 0 |
| GIGYF2 | low | 0 | 0 | 0 | 0 | 0 | 0 | 0 | 0 | 0 | 0 | 0 | 0 |
| TMEM230 | low | 1 | 0 | 0 | 0 | 0 | 0 | 0 | 0 | 1 | 0 | 0 | 0 |
| UCHL1 | low | 0 | 0 | 0 | 0 | 0 | 0 | 0 | 0 | 0 | 0 | 0 | 0 |

PD: Parkinson's Disease
AAC: African American AFR: African AJ: Ashkenazi Jew AMR: Admixed American/Latin American CAH: Complex Admixture History CAS: Central Asian EAS: East Asian EUR: European FIN: Finnish MDE: Middle Eastern SAS: South Asian

A breakdown of all pathogenic variants identified across the different ancestries shows the number of variants unique to specific ancestries is quite small but biased towards the EUR ancestry, which has 11 unique variants not observed in any other ancestry (Supplementary Table 5). Of further relevance is the fact that the SAS ancestry group had no pathogenic variants present within the PD genes of interest. Interestingly, each of the pathogenic variants within *PRKN* were found to be present in only one ancestry group.

## Discussion

With the increase in prevalence of PD (20), it is important to enhance our understanding of the genetic influence of pathogenic variants across ancestrally diverse populations. Through our analysis of cluster plots and allele frequencies, we sought to evaluate the effectiveness of the NBA for genotyping pathogenic variants.

From the cluster plots, we are able to discern that the NBA is an effective platform for studying PD across diverse populations. The number of NCs for pathogenic variants were high in genes that are historically difficult to genotype, and were also higher on average in variants that were classified as “bad”. High quality genotyping is vital for accurately identifying pathogenic variants, yet it is often assumed to be adequate. For example, the pathogenic gene *GBA1* is located in a region with a complex sequence and is prone to copy number variants, making it prone to misalignment or errors during genotyping (21). Our study revealed that of the 14 typed pathogenic variants within *GBA1*, 2 were classified as “medium” and 4 were classified as “bad” based on their cluster plots. One of the “bad” pathogenic variants within *GBA1* had 91 NCs, further confirming the difficulty in genotyping SNPs in this gene. Though the NBA is an advanced tool for genotype analysis for PD and other NDDs, the existence of “medium” and “bad” variants underscores the importance in verifying the quality of genotypes when using them for further studies.

We found more pathogenic variants in genes classified as having a “very high” probability of causing PD by Blauwendraat et al. compared to genes classified as “low” probability of causing PD. Many of the genes that were classified as having “low” confidence of causing PD had no pathogenic variants present in our data. It must be noted, however, that the NBA was designed to capture the pathogenic variants in established PD genes, which could contribute to why more pathogenic variants were found in genes with a “very high” influence on the risk for PD. Interestingly, *SNCA, FBXO7, VPS35*, and *PARK7* were found to have no pathogenic variants present in our data even though they have a “very high” confidence in contributing to PD. This may be contributed to the fact that the pathogenic variants within these genes are extremely rare (i.e. not present in our data) as is the case with the pathogenic variants in *SNCA* (PMID: 33307186). Even gnomAD had no pathogenic variants for PD reported in *SNCA* and all pathogenic variants in *FBXO7* in gnomAD had an allele frequency of >=1.30E-5. It is also possible that our understanding of the pathogenicity of these variants has changed over time, and the ClinVar database needs to be updated to reflect these changes.

Comparing across ancestries, it is noteworthy that certain pathogenic variants are present only in specific ancestries. For example, the single annotated pathogenic variant in *LRRK2* (rs33939927) is present in only the CAS, EUR, and MDE populations. The single annotated pathogenic variant in *TMEM230*, a gene classified as having a “low” confidence in contributing to PD by Blauwendraat et al., is only present in EUR. This may be explained by the fact that many genetic studies identifying pathogenic genes for PD have historically focused on samples of European ancestry, which may skew the importance of these genes and not represent its global significance. The low frequency of variants in ancestries other than EUR may also be due to the fact that the ancestry sample size is notably smaller than the EUR ancestry in GP2. It is also interesting to note that different pathogenic variants within a gene are present in unique ancestries, such as in *GBA1* where all of its pathogenic variants are present in only one or two ancestries. This underscores the importance in conducting research on ancestrally diverse samples, corroborating previous claims of the lack of population diversity in PD research (13).

Some limitations of our report include the fact that many of the pathogenic variants were imputed using the TOPMed Imputation Server (22) instead of being directly typed from the NBA. Because they were not typed by the NBA, we were unable to create cluster plots for many of the pathogenic variants. The categorization of the NBA cluster plots was done by manually evaluating the visualizations, which can also lead to bias in the classifications of “medium” variants. The number of samples is also not consistent across ancestries causing some ancestral groups to have more power than others. Although our study included a variety of ancestral backgrounds to combat the lack of diversity in the study of PD genes, our largest demographic was still EUR with 38,839 samples. Finally, our reliance on the ClinVar database to denote variants as “Pathogenic” limits our analysis to variants within the scope of the ClinVar annotations. ClinVar can also be biased as it labels variants as “Pathogenic” from studies primarily published in European populations.

GP2’s initiative to provide a more comprehensive, diverse population for studying PD has clinical and practical implications in the study of PD genetics. Our study reveals that conventional PD genes that were previously discovered in cohorts of predominantly European ancestry may not have the same implications in non-European populations. Additionally, our evaluation of the NBA’s genotyping success underscores its effectiveness in identifying these variants. This work highlights the critical role of GP2 in enhancing our understanding of PD genetics across diverse populations and underscores NBA’s utility in robustly detecting pathogenic variants.

## Data Availability

Data (DOI 10.5281/zenodo.10962119, release 7) used in the preparation of this article were obtained from the Global Parkinson's Genetics Program (GP2). GP2 is funded by the Aligning Science Across Parkinson's (ASAP) initiative and implemented by The Michael J. Fox Foundation for Parkinson's Research (https://gp2.org). For a complete list of GP2 members see https://gp2.org.
Data used in the preparation of this article were obtained from the Accelerating Medicine Partnership (AMP) Parkinson's Disease (AMP PD) Knowledge Platform. For up-to-date information on the study, visit https://www.amp-pd.org. The AMP PD program is a public-private partnership managed by the Foundation for the National Institutes of Health and funded by the National Institute of Neurological Disorders and Stroke (NINDS) in partnership with the Aligning Science Across Parkinson's (ASAP) initiative; Celgene Corporation, a subsidiary of Bristol-Myers Squibb Company; GlaxoSmithKline plc (GSK); The Michael J. Fox Foundation for Parkinson's Research; Pfizer Inc.; AbbVie Inc.; Sanofi US Services Inc.; and Verily Life Sciences. ACCELERATING MEDICINES PARTNERSHIP and AMP are registered service marks of the U.S. Department of Health and Human Services.

https://github.com/GP2code/pathogenic_pd_variants_clusters

## Acknowledgements

Data (DOI 10.5281/zenodo.10962119, release 7) used in the preparation of this article were obtained from the Global Parkinson’s Genetics Program (GP2). GP2 is funded by the Aligning Science Across Parkinson’s (ASAP) initiative and implemented by The Michael J. Fox Foundation for Parkinson’s Research (https://gp2.org). For a complete list of GP2 members see https://gp2.org.

Data used in the preparation of this article were obtained from the Accelerating Medicine Partnership® (AMP®) Parkinson’s Disease (AMP PD) Knowledge Platform. For up-to-date information on the study, visit https://www.amp-pd.org. The AMP® PD program is a public-private partnership managed by the Foundation for the National Institutes of Health and funded by the National Institute of Neurological Disorders and Stroke (NINDS) in partnership with the Aligning Science Across Parkinson’s (ASAP) initiative; Celgene Corporation, a subsidiary of Bristol-Myers Squibb Company; GlaxoSmithKline plc (GSK); The Michael J. Fox Foundation for Parkinson’s Research; Pfizer Inc.; AbbVie Inc.; Sanofi US Services Inc.; and Verily Life Sciences. ACCELERATING MEDICINES PARTNERSHIP and AMP are registered service marks of the U.S. Department of Health and Human Services.

## Notes

**Financial Disclosures/Conflict of Interest:** M.J.K and H.L.’s participation in this project was part of a competitive contract awarded to DataTecnica LLC by the National Institutes of Health to support open science research. V.P. serves as an Editorial Board Member for Nature Portfolio.

### Competing Interest Statement

M.J.K and H.L.'s participation in this project was part of a competitive contract awarded to DataTecnica LLC by the National Institutes of Health to support open science research. V.P. serves as an Editorial Board Member for Nature Portfolio.

### Funding Statement

This research was supported in part by the Intramural Research Program of the NIH, National Institute on Aging (NIA), National Institutes of Health, Department of Health and Human Services; project number ZO1 AG000534, as well as the National Institute of Neurological Disorders and Stroke.

### Author Declarations

Data (DOI 10.5281/zenodo.10962119, release 7) used in the preparation of this article were obtained from the Global Parkinson's Genetics Program (GP2). GP2 is funded by the Aligning Science Across Parkinson's (ASAP) initiative and implemented by The Michael J. Fox Foundation for Parkinson's Research (https://gp2.org). For a complete list of GP2 members see https://gp2.org. Data used in the preparation of this article were obtained from the Accelerating Medicine Partnership (AMP) Parkinson's Disease (AMP PD) Knowledge Platform. For up-to-date information on the study, visit https://www.amp-pd.org. The AMP PD program is a public-private partnership managed by the Foundation for the National Institutes of Health and funded by the National Institute of Neurological Disorders and Stroke (NINDS) in partnership with the Aligning Science Across Parkinson's (ASAP) initiative; Celgene Corporation, a subsidiary of Bristol-Myers Squibb Company; GlaxoSmithKline plc (GSK); The Michael J. Fox Foundation for Parkinson's Research; Pfizer Inc.; AbbVie Inc.; Sanofi US Services Inc.; and Verily Life Sciences. ACCELERATING MEDICINES PARTNERSHIP and AMP are registered service marks of the U.S. Department of Health and Human Services. The datasets used in this study were individual-level data, and that individual-level data was de-identified prior to its use in this study.

## REFERENCES

1. Chai C, Lim KL. Genetic insights into sporadic Parkinson’s disease pathogenesis. Curr Genomics. 2013 Dec;14(8):486–501.

2. Lim SY, Klein C. Parkinson’s Disease is Predominantly a Genetic Disease. J Parkinsons Dis. 2024;14(3):467–82.

3. Dorsey ER, Bloem BR. Parkinson’s Disease Is Predominantly an Environmental Disease. J Parkinsons Dis. 2024;14(3):451–65.

4. Wood-Kaczmar A, Gandhi S, Wood NW. Understanding the molecular causes of Parkinson’s disease. Trends Mol Med. 2006 Nov;12(11):521–8.

5. Nuytemans K, Theuns J, Cruts M, Van Broeckhoven C. Genetic etiology of Parkinson disease associated with mutations in the SNCA, PARK2, PINK1, PARK7, and LRRK2 genes: a mutation update. Hum Mutat. 2010 Jul;31(7):763–80.

6. Oczkowska A, Kozubski W, Lianeri M, Dorszewska J. Mutations in PRKN and SNCA Genes Important for the Progress of Parkinson’s Disease. Curr Genomics. 2013 Dec;14(8):502–17.

7. Klein C, Westenberger A. Genetics of Parkinson’s disease. Cold Spring Harb Perspect Med. 2012 Jan;2(1):a008888.

8. Nalls MA, Blauwendraat C, Vallerga CL, Heilbron K, Bandres-Ciga S, Chang D, et al. Identification of novel risk loci, causal insights, and heritable risk for Parkinson’s disease: a meta-analysis of genome-wide association studies. Lancet Neurol. 2019 Dec;18(12):1091–102.

9. Foo JN, Chew EGY, Chung SJ, Peng R, Blauwendraat C, Nalls MA, et al. Identification of Risk Loci for Parkinson Disease in Asians and Comparison of Risk Between Asians and Europeans: A Genome-Wide Association Study. JAMA Neurol. 2020 Jun 1;77(6):746–54.

10. Loesch DP, Horimoto ARVR, Heilbron K, Sarihan EI, Inca-Martinez M, Mason E, et al. Characterizing the Genetic Architecture of Parkinson’s Disease in Latinos. Ann Neurol. 2021 Sep;90(3):353–65.

11. Rizig M, Bandres-Ciga S, Makarious MB, Ojo OO, Crea PW, Abiodun OV, et al. Identification of genetic risk loci and causal insights associated with Parkinson’s disease in African and African admixed populations: a genome-wide association study. Lancet Neurol. 2023 Nov;22(11):1015–25.

12. Blauwendraat C, Nalls MA, Singleton AB. The genetic architecture of Parkinson’s disease. Lancet Neurol. 2020 Feb;19(2):170–8.

13. Schumacher-Schuh AF, Bieger A, Okunoye O, Mok KY, Lim SY, Bardien S, et al. Underrepresented Populations in Parkinson’s Genetics Research: Current Landscape and Future Directions. Mov Disord. 2022 Aug;37(8):1593–604.

14. Bandres-Ciga S, Faghri F, Majounie E, Koretsky MJ, Kim J, Levine KS, et al. NeuroBooster Array: A Genome-Wide Genotyping Platform to Study Neurological Disorders Across Diverse Populations. medRxiv [Internet]. 2023 Nov 14; Available from: 10.1101/2023.11.06.23298176

15. Vitale D, Koretsky M, Kuznetsov N, Hong S, Martin J, James M, et al. GenoTools: An Open-Source Python Package for Efficient Genotype Data Quality Control and Analysis. bioRxiv [Internet]. 2024 Jul 3; Available from: 10.1101/2024.03.26.586362

16. Wang K, Li M, Hakonarson H. ANNOVAR: functional annotation of genetic variants from high-throughput sequencing data. Nucleic Acids Res. 2010 Sep;38(16):e164.

17. Chang CC, Chow CC, Tellier LC, Vattikuti S, Purcell SM, Lee JJ. Second-generation PLINK: rising to the challenge of larger and richer datasets. Gigascience. 2015 Feb 25;4:7.

18. Tindall EA, Petersen DC, Nikolaysen S, Miller W, Schuster SC, Hayes VM. Interpretation of custom designed Illumina genotype cluster plots for targeted association studies and next-generation sequence validation. BMC Res Notes. 2010 Feb 22;3:39.

19. Staaf J, Vallon-Christersson J, Lindgren D, Juliusson G, Rosenquist R, Höglund M, et al. Normalization of Illumina Infinium whole-genome SNP data improves copy number estimates and allelic intensity ratios. BMC Bioinformatics. 2008 Oct 2;9:409.

20. Pringsheim T, Jette N, Frolkis A, Steeves TDL. The prevalence of Parkinson’s disease: a systematic review and meta-analysis. Mov Disord. 2014 Nov;29(13):1583–90.

21. Woo EG, Tayebi N, Sidransky E. Next-Generation Sequencing Analysis of : The Challenge of Detecting Complex Recombinant Alleles. Front Genet. 2021 Jun 21;12:684067.

22. Das S, Forer L, Schönherr S, Sidore C, Locke AE, Kwong A, et al. Next-generation genotype imputation service and methods. Nat Genet. 2016 Oct;48(10):1284–7.

