## Supplementary Figure 1 for "Parkinson’s Disease Pathogenic Variants: Cross-Ancestry Analysis and Microarray Data Validation"

**Selected example cluster plots**

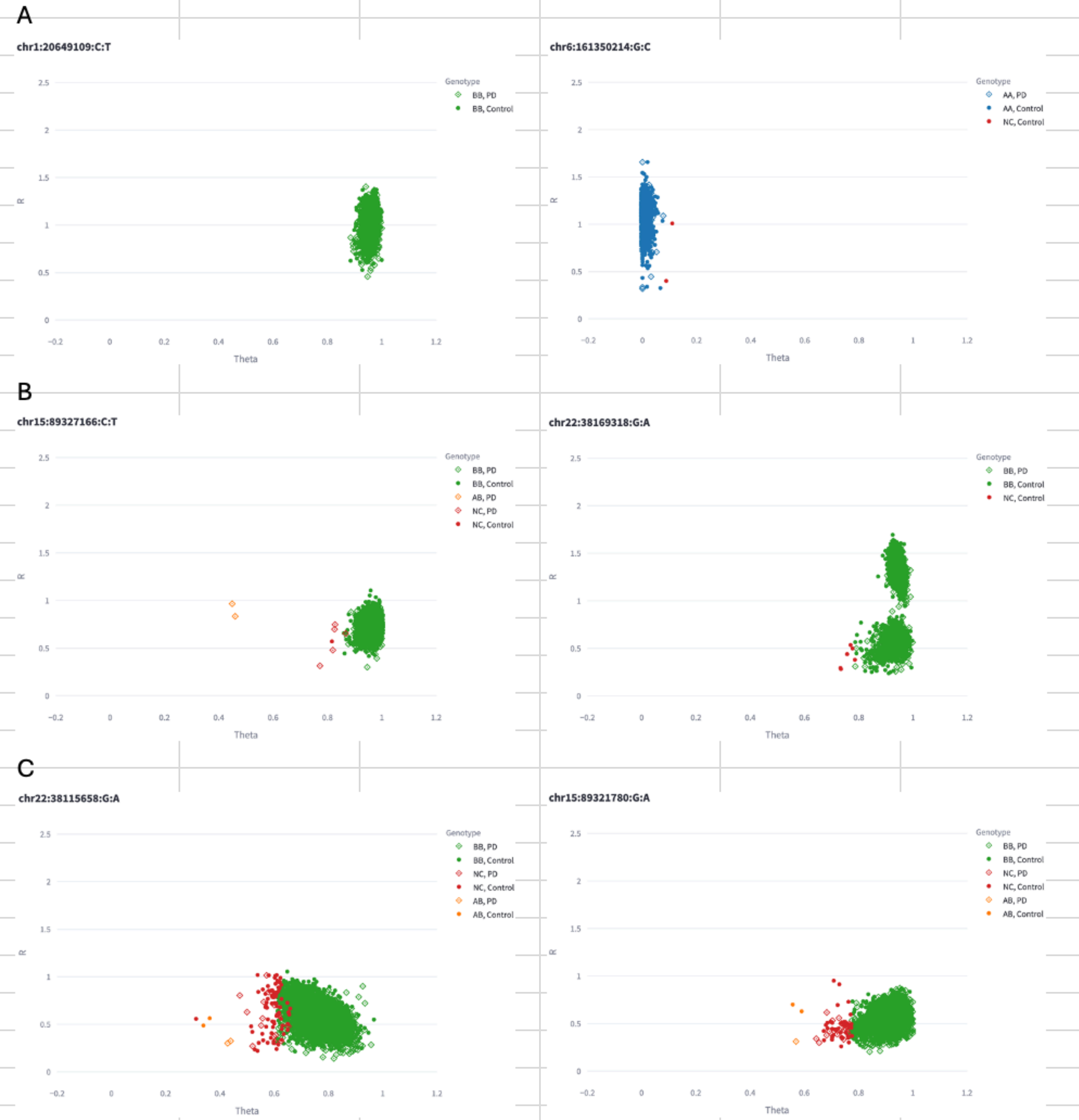


Supplementary Figure 1. Examples of cluster plots per each classification

A. Examples of "good" (good clusters, limited no calls)
B. Examples of "medium" (some bleeding between clusters, some no calls)
C. Examples of "bad" (poor clusters, many no calls)
