## Supplementary Table 1 for "Parkinson’s Disease Pathogenic Variants: Cross-Ancestry Analysis and Microarray Data Validation"

**Demographics Overview**

| Ancestry | Total | | | Cases ("PD") | | | Control ("Control") | | | Case ("Other") | | |
|---|---|---|---|---|---|---|---|---|---|---|---|---|
| | n | age(mean) | age(sd) | n | age(mean) | age(sd) | n | age(mean) | age(sd) | n | age(mean) | age(sd) |
| AAC | 1,111 | 65.78 | 10.45 | 285 | 65.45 | 11.07 | 801 | 65.84 | 10.27 | 25 | 67.34 | 10.32 |
| AFR | 2,643 | 63.56 | 14.81 | 942 | 63.26 | 12.14 | 1,679 | 65.59 | 15.36 | 22 | 66.87 | 10.25 |
| AJ | 2,655 | 67.96 | 10.55 | 1,292 | 70.04 | 9.89 | 411 | 67.81 | 9.88 | 952 | 65.03 | 10.96 |
| AMR | 646 | 61.64 | 11.93 | 458 | 61.65 | 12.71 | 155 | 61.19 | 9.45 | 33 | 63.90 | 8.95 |
| CAH | 851 | 55.44 | 17.17 | 525 | 61.06 | 12.73 | 302 | 46.50 | 19.39 | 24 | 63.98 | 9.48 |
| CAS | 903 | 58.92 | 9.72 | 552 | 61.02 | 10.68 | 343 | 55.05 | 5.75 | 8 | 60.31 | 15.98 |
| EAS | 5,167 | 65.01 | 10.93 | 2,662 | 67.51 | 10.09 | 2,461 | 62.40 | 11.15 | 44 | 70.88 | 8.94 |
| EUR | 38,839 | 65.54 | 11.27 | 21,198 | 66.74 | 10.89 | 9,214 | 62.11 | 13.16 | 8,427 | 65.80 | 9.62 |
| FIN | 114 | 65.91 | 11.34 | 98 | 63.95 | 10.95 | 8 | 76.15 | 11.78 | 8 | 71.25 | 6.75 |
| MDE | 581 | 59.37 | 12.00 | 311 | 64.14 | 12.41 | 225 | 55.45 | 9.69 | 45 | 58.67 | 13.74 |
| SAS | 635 | 59.74 | 15.02 | 387 | 62.94 | 12.50 | 222 | 54.95 | 16.84 | 26 | 73.40 | 5.55 |
| Total | 54,145 | 65.12 | 11.64 | 28,710 | 66.54 | 11.03 | 15,821 | 61.83 | 13.37 | 9,614 | 65.7 | 9.82 |

PD: Parkinson's Disease

AAC: African American
AFR: African
AJ: Ashkenazi Jew
AMR: Admixed American/Latin American
CAH: Complex Admixture History
CAS: Central Asian
EAS: East Asian
EUR: European
FIN: Finnish
MDE: Middle Eastern
SAS: South Asian
