## Supplementary Table 2 for "Parkinson’s Disease Pathogenic Variants: Cross-Ancestry Analysis and Microarray Data Validation"

**ClinVar Annotations for the Pathogenic Variants within Established PD Genes**

| Chr | Start | End | Ref | Alt | Func.refGene | Gene.refGene | GeneDetail.refGene | ExonicFunc.refGene | AAChange.refGene | avsnp151 | CLNALLELEID | CLNDN | CLNDISDB | CLNREVSTAT | CLNSIG | SIFT_score | SIFT converted rankscore | SIFT_pred |
|---|---|---|---|---|---|---|---|---|---|---|---|---|---|---|---|---|---|---|
| chr12 | 40310434 | 40310434 | C | T | exonic | LRRK2 | . | nonsynonymous SNV | LRRK2:NM_198578:exon31:c.C4321T:p.R1441C | rs33939927 | 16977 | not_provided\|Autosomal_dominant_Parkinson_disease_8 | MedGen:C3661900\|MONDO:MOND | criteria_provided,_multiple_submitters,_no_conflicts | Pathogenic | 0.117 | 0.282 | T |
| chr15 | 89319225 | 89319225 | T | A | intronic | POLG | . | . | . | rs778573169 | 580246 | not_provided\|Inborn_genetic_diseases\|Progressive_sclerosing_poliodystrophy | MedGen:C3661900\|MeSH:D030342, | criteria_provided,_multiple_submitters,_no_conflicts | Pathogenic | . | . | . |
| chr15 | 89321780 | 89321780 | G | A | exonic | POLG | . | nonsynonymous SNV | POLG:NM_001126131:exon16:c.C2554T:p.R852C,<br>POLG:NM_002693:exon16:c.C2554T:p.R852C | rs144500145 | 202960 | Inborn_genetic_diseases\|POLG-Related_Spectrum_Disorders\|Intellectual_disability\|not_provided\| | MeSH:D030342,MedGen:C0950123\| | criteria_provided,_multiple_submitters,_no_conflicts | Pathogenic | 0 | 0.912 | D |
| chr15 | 89321792 | 89321792 | C | T | exonic | POLG | . | nonsynonymous SNV | POLG:NM_001126131:exon16:c.G2542A:p.G848S,<br>POLG:NM_002693:exon16:c.G2542A:p.G848S | rs113994098 | 28541 | Progressive_sclerosing_poliodystrophy\|Mitochondrial_DNA_depletion_syndrome_4b\|Sensory_atax | MONDO:MONDO:0008758,MedGen: | criteria_provided,_multiple_submitters,_no_conflicts | Pathogenic | 0 | 0.912 | D |
| chr15 | 89325610 | 89325610 | G | A | exonic | POLG | . | nonsynonymous SNV | POLG:NM_001126131:exon10:c.C1789T:p.R597W,<br>POLG:NM_002693:exon10:c.C1789T:p.R597W | rs139717885 | 374507 | Progressive_sclerosing_poliodystrophy\|not_provided\|Sensory_ataxic_neuropathy,_dysarthria,_and_ | MONDO:MONDO:0008758,MedGen: | criteria_provided,_multiple_submitters,_no_conflicts | Pathogenic | 0.002 | 0.721 | D |
| chr15 | 89325679 | 89325679 | G | A | exonic | POLG | . | nonsynonymous SNV | POLG:NM_001126131:exon10:c.C1720T:p.R574W,<br>POLG:NM_002693:exon10:c.C1720T:p.R574W | rs774474723 | 1438803 | Progressive_sclerosing_poliodystrophy\|not_provided | MONDO:MONDO:0008758,MedGen: | criteria_provided,_multiple_submitters,_no_conflicts | Pathogenic | 0.001 | 0.784 | D |
| chr15 | 89327166 | 89327166 | C | T | splicing | POLG | NM_001126131:exon7:c.1433+1G>A;<br>NM_002693:exon7:c.1433+1G>A | . | . | rs771623994 | 264590 | Progressive_sclerosing_poliodystrophy\|POLG-related_disorder\|not_provided\|Progressive_external_ | MONDO:MONDO:0008758,MedGen: | criteria_provided,_multiple_submitters,_no_conflicts | Pathogenic | . | . | . |
| chr15 | 89327201 | 89327201 | C | T | exonic | POLG | . | nonsynonymous SNV | POLG:NM_001126131:exon7:c.G1399A:p.A467T,<br>POLG:NM_002693:exon7:c.G1399A:p.A467T | rs113994095 | 28535 | Progressive_sclerosing_poliodystrophy\|Mitochondrial_DNA_depletion_syndrome_4b\|Sensory_atax | MONDO:MONDO:0008758,MedGen: | criteria_provided,_multiple_submitters,_no_conflicts | Pathogenic | 0.003 | 0.682 | D |
| chr15 | 89329041 | 89329041 | G | A | exonic | POLG | . | nonsynonymous SNV | POLG:NM_001126131:exon4:c.C925T:p.R309C,<br>POLG:NM_002693:exon4:c.C925T:p.R309C | rs886041592 | 264743 | not_provided\|Progressive_sclerosing_poliodystrophy\|Progressive_external_ophthalmoplegia_with_ | MedGen:C3661900\|MONDO:MOND | criteria_provided,_multiple_submitters,_no_conflicts | Pathogenic | 0 | 0.912 | D |
| chr1 | 155235196 | 155235196 | G | A | exonic | GBA | . | nonsynonymous SNV | GBA:NM_001171811:exon9:c.C1243T:p.R415C,<br>GBA:NM_001171812:exon9:c.C1357T:p.R453C,<br>GBA:NM_000157:exon10:c.C1504T:p.R502C,<br>GBA:NM_001005741:exon11:c.C1504T:p.R502C,<br>GBA:NM_001005742:exon11:c.C1504T:p.R502C | rs80356771 | 19334 | Parkinson_disease,_late-onset\|Gaucher_disease-ophthalmoplegia-cardiovascular_calcification_sy | MONDO:MONDO:0008199,MedGen: | criteria_provided,_multiple_submitters,_no_conflicts | Pathogenic | 0.001 | 0.784 | D |
| chr1 | 155236295 | 155236295 | G | A | exonic | GBA | . | nonsynonymous SNV | GBA:NM_001171811:exon7:c.C913T:p.R305W,<br>GBA:NM_001171812:exon7:c.C1027T:p.R343W,<br>GBA:NM_000157:exon8:c.C1174T:p.R392W,<br>GBA:NM_001005741:exon9:c.C1174T:p.R392W,<br>GBA:NM_001005742:exon9:c.C1174T:p.R392W | rs121908308 | 801582 | Gaucher_disease_type_I\|Gaucher_disease_type_II\|Gaucher_disease_type_III\|Gaucher_disease-o | MONDO:MONDO:0009265,MedGen: | criteria_provided,_single_submitter | Pathogenic | 0.024 | 0.501 | D |
| chr1 | 155237453 | 155237453 | C | T | exonic | GBA | . | nonsynonymous SNV | GBA:NM_001171811:exon6:c.G626A:p.R209Q,<br>GBA:NM_001171812:exon6:c.G740A:p.R247Q,<br>GBA:NM_000157:exon7:c.G887A:p.R296Q,<br>GBA:NM_001005741:exon8:c.G887A:p.R296Q,<br>GBA:NM_001005742:exon8:c.G887A:p.R296Q | rs78973108 | 19367 | Parkinson_disease,_late-onset\|Gaucher_disease-ophthalmoplegia-cardiovascular_calcification_sy | MONDO:MONDO:0008199,MedGen: | criteria_provided,_multiple_submitters,_no_conflicts | Pathogenic | 0.009 | 0.614 | D |
| chr1 | 155238206 | 155238206 | A | C | exonic | GBA | . | nonsynonymous SNV | GBA:NM_001171811:exon5:c.T428G:p.V143G,<br>GBA:NM_001171812:exon5:c.T542G:p.V181G,<br>GBA:NM_000157:exon6:c.T689G:p.V230G,<br>GBA:NM_001005741:exon7:c.T689G:p.V230G,<br>GBA:NM_001005742:exon7:c.T689G:p.V230G | rs381427 | 916795 | Gaucher_disease | MONDO:MONDO:0018150,MedGen: | criteria_provided,_single_submitter | Pathogenic | 0.01 | 0.564 | D |
| chr1 | 155238215 | 155238215 | T | C | exonic | GBA | . | nonsynonymous SNV | GBA:NM_001171811:exon5:c.A419G:p.N140S,<br>GBA:NM_001171812:exon5:c.A533G:p.N178S,<br>GBA:NM_000157:exon6:c.A680G:p.N227S,<br>GBA:NM_001005741:exon7:c.A680G:p.N227S,<br>GBA:NM_001005742:exon7:c.A680G:p.N227S | rs364897 | 19353 | Gaucher_disease-ophthalmoplegia-cardiovascular_calcification_syndrome\|Gaucher_disease_type_ | MONDO:MONDO:0009268,MedGen: | criteria_provided,_multiple_submitters,_no_conflicts | Pathogenic | 0.241 | 0.191 | T |
| chr1 | 155238630 | 155238630 | G | A | exonic | GBA | . | nonsynonymous SNV | GBA:NM_001171811:exon4:c.C214T:p.R72W,<br>GBA:NM_001171812:exon4:c.C328T:p.R110W,<br>GBA:NM_000157:exon5:c.C475T:p.R159W,<br>GBA:NM_001005741:exon6:c.C475T:p.R159W,<br>GBA:NM_001005742:exon6:c.C475T:p.R159W | rs439898 | 76478 | Gaucher_disease\|not_provided | MONDO:MONDO:0018150,MedGen: | criteria_provided,_multiple_submitters,_no_conflicts | Pathogenic | 0 | 0.912 | D |
| chr1 | 155239934 | 155239934 | G | A | exonic | GBA | . | nonsynonymous SNV | GBA:NM_000157:exon3:c.C259T:p.R87W,<br>GBA:NM_001171812:exon3:c.C259T:p.R87W,<br>GBA:NM_001005741:exon4:c.C259T:p.R87W,<br>GBA:NM_001005742:exon4:c.C259T:p.R87W | rs1141814 | 19360 | Parkinson_disease,_late-onset\|Gaucher_disease-ophthalmoplegia-cardiovascular_calcification_sy | MONDO:MONDO:0008199,MedGen: | criteria_provided,_multiple_submitters,_no_conflicts | Pathogenic | 0 | 0.912 | D |
| chr1 | 16988455 | 16988455 | C | T | exonic | ATP13A2 | . | nonsynonymous SNV | ATP13A2:NM_001141974:exon23:c.G2497A:p.G833R<br>ATP13A2:NM_001141973:exon24:c.G2614A:p.G872R<br>ATP13A2:NM_022089:exon24:c.G2629A:p.G877R | rs144701072 | 77001 | Kufor-Rakeb_syndrome | MONDO:MONDO:0011706,MedGen: | no_assertion_criteria_provided | Pathogenic | 0 | 0.912 | D |
| chr1 | 16989961 | 16989961 | G | A | exonic | ATP13A2 | . | stopgain | ATP13A2:NM_001141973:exon22:c.C2440T:p.R814X,<br>ATP13A2:NM_022089:exon22:c.C2455T:p.R819X | rs866035312 | 493540 | not_provided\|Autosomal_recessive_spastic_paraplegia_type_78\|Kufor-Rakeb_syndrome | MedGen:C3661900\|MONDO:MOND | criteria_provided,_multiple_submitters,_no_conflicts | Pathogenic | . | . | . |
| chr1 | 16992345 | 16992345 | G | A | exonic | ATP13A2 | . | stopgain | ATP13A2:NM_001141973:exon18:c.C1888T:p.Q630X,<br>ATP13A2:NM_001141974:exon18:c.C1888T:p.Q630X,<br>ATP13A2:NM_022089:exon18:c.C1903T:p.Q635X | rs773246271 | 447471 | Kufor-Rakeb_syndrome\|Autosomal_recessive_spastic_paraplegia_type_78\|not_provided | MONDO:MONDO:0011706,MedGen: | criteria_provided,_multiple_submitters,_no_conflicts | Pathogenic | . | . | . |
| chr1 | 16996059 | 16996059 | G | A | exonic | ATP13A2 | . | stopgain | ATP13A2:NM_001141973:exon15:c.C1444T:p.R482X,<br>ATP13A2:NM_001141974:exon15:c.C1444T:p.R482X,<br>ATP13A2:NM_022089:exon15:c.C1459T:p.R487X | rs1303653650 | 511191 | Inborn_genetic_diseases\|Kufor-Rakeb_syndrome\|Autosomal_recessive_spastic_paraplegia_type_ | MeSH:D030342,MedGen:C0950123\| | criteria_provided,_multiple_submitters,_no_conflicts | Pathogenic | . | . | . |
| chr1 | 17005450 | 17005450 | C | T | exonic | ATP13A2 | . | stopgain | ATP13A2:NM_001141973:exon3:c.G212A:p.W71X,<br>ATP13A2:NM_001141974:exon3:c.G212A:p.W71X,<br>ATP13A2:NM_022089:exon3:c.G212A:p.W71X | rs373607247 | 425322 | not_provided\|Inborn_genetic_diseases | MedGen:CN517202\|MeSH:D030342, | criteria_provided,_multiple_submitters,_no_conflicts | Pathogenic | . | . | . |
| chr1 | 20645640 | 20645640 | T | C | exonic | PINK1 | . | nonsynonymous SNV | PINK1:NM_032409:exon5:c.T1040C:p.L347P | rs28940285 | 17447 | Autosomal_recessive_early-onset_Parkinson_disease_6\|not_provided | MONDO:MONDO:0011613,MedGen: | criteria_provided,_multiple_submitters,_no_conflicts | Pathogenic | 0 | 0.912 | D |
| chr1 | 20649109 | 20649109 | C | T | exonic | PINK1 | . | stopgain | PINK1:NM_032409:exon7:c.C1366T:p.Q456X | rs45539432 | 17454 | not_provided\|Autosomal_recessive_early-onset_Parkinson_disease_6 | MedGen:C3661900\|MONDO:MOND | criteria_provided,_multiple_submitters,_no_conflicts | Pathogenic | . | . | . |
| chr1 | 20649217 | 20649217 | C | T | exonic | PINK1 | . | stopgain | PINK1:NM_032409:exon7:c.C1474T:p.R492X | rs34208370 | 425331 | Autosomal_recessive_early-onset_Parkinson_disease_6\|not_provided | MONDO:MONDO:0011613,MedGen: | criteria_provided,_multiple_submitters,_no_conflicts | Pathogenic | . | . | . |
| chr20 | 5100921 | 5100921 | C | A | exonic | TMEM230 | . | nonsynonymous SNV | TMEM230:NM_001009925:exon4:c.G233T:p.R78L,<br>TMEM230:NM_001330985:exon4:c.G233T:p.R78L,<br>TMEM230:NM_001330986:exon4:c.G233T:p.R78L,<br>TMEM230:NM_014145:exon4:c.G233T:p.R78L,<br>TMEM230:NM_001009923:exon5:c.G422T:p.R141L,<br>TMEM230:NM_001009924:exon5:c.G233T:p.R78L,<br>TMEM230:NM_001330984:exon5:c.G233T:p.R78L | rs764786986 | 244095 | not_provided | MedGen:C3661900 | criteria_provided,_single_submitter | Pathogenic | 0.005 | 0.654 | D |
| chr21 | 32726907 | 32726907 | C | A | exonic | SYNJ1 | . | stopgain | SYNJ1:NM_003895:exon2:c.G106T:p.E36X | rs1040540690 | 1393547 | Developmental_and_epileptic_encephalopathy,_53\|Early-onset_Parkinson_disease_20 | MONDO:MONDO:0033362,MedGen: | criteria_provided,_single_submitter | Pathogenic | . | . | . |
| chr22 | 38115658 | 38115658 | G | A | exonic | PLA2G6 | . | stopgain | PLA2G6:NM_001004426:exon13:c.C1741T:p.R581X,<br>PLA2G6:NM_001199562:exon13:c.C1741T:p.R581X,<br>PLA2G6:NM_001349865:exon13:c.C1741T:p.R581X,<br>PLA2G6:NM_001349866:exon13:c.C1741T:p.R581X,<br>PLA2G6:NM_001349868:exon13:c.C1225T:p.R409X,<br>PLA2G6:NM_001349864:exon14:c.C1903T:p.R635X,<br>PLA2G6:NM_001349869:exon14:c.C1207T:p.R403X,<br>PLA2G6:NM_003560:exon14:c.C1903T:p.R635X,<br>PLA2G6:NM_001349867:exon15:c.C1369T:p.R457X | rs587784339 | 169779 | Infantile_neuroaxonal_dystrophy\|PLA2G6-associated_neurodegeneration\|not_provided\|Iron_accum | MONDO:MONDO:0024457,MedGen: | criteria_provided,_multiple_submitters,_no_conflicts | Pathogenic | . | . | . |
| chr22 | 38132917 | 38132917 | C | A | exonic | PLA2G6 | . | nonsynonymous SNV | PLA2G6:NM_001349868:exon6:c.G313T:p.D105Y,<br>PLA2G6:NM_001004426:exon7:c.G991T:p.D331Y,<br>PLA2G6:NM_001199562:exon7:c.G991T:p.D331Y,<br>PLA2G6:NM_001349864:exon7:c.G991T:p.D331Y,<br>PLA2G6:NM_001349865:exon7:c.G991T:p.D331Y,<br>PLA2G6:NM_001349866:exon7:c.G991T:p.D331Y,<br>PLA2G6:NM_003560:exon7:c.G991T:p.D331Y,<br>PLA2G6:NM_001349867:exon8:c.G457T:p.D153Y,<br>PLA2G6:NM_001349869:exon8:c.G457T:p.D153Y | rs199935023 | 39328 | Infantile_neuroaxonal_dystrophy\|Autosomal_recessive_Parkinson_disease_14\|PLA2G6-associated | MONDO:MONDO:0024457,MedGen: | criteria_provided,_multiple_submitters,_no_conflicts | Pathogenic | 0.003 | 0.682 | D |
| chr22 | 38169318 | 38169318 | G | A | exonic | PLA2G6 | . | stopgain | PLA2G6:NM_001004426:exon2:c.C109T:p.R37X,<br>PLA2G6:NM_001199562:exon2:c.C109T:p.R37X,<br>PLA2G6:NM_001349864:exon2:c.C109T:p.R37X,<br>PLA2G6:NM_001349865:exon2:c.C109T:p.R37X,<br>PLA2G6:NM_001349866:exon2:c.C109T:p.R37X,<br>PLA2G6:NM_003560:exon2:c.C109T:p.R37X | rs200075782 | 39327 | Iron_accumulation_in_brain\|Neurodegeneration_with_brain_iron_accumulation_2B\|Infantile_neuroa | Human_Phenotype_Ontology:HP:001 | criteria_provided,_multiple_submitters,_no_conflicts | Pathogenic | . | . | . |
| chr6 | 161350214 | 161350214 | G | C | intronic | PRKN | . | . | . | rs765860776 | 1867959 | not_provided | MedGen:C3661900 | criteria_provided,_single_submitter | Pathogenic | . | . | . |
| chr6 | 161785793 | 161785793 | C | G | exonic | PRKN | . | nonsynonymous SNV | PRKN:NM_013988:exon4:c.G403C:p.G135R,<br>PRKN:NM_013987:exon6:c.G766C:p.G256R,<br>PRKN:NM_004562:exon7:c.G850C:p.G284R | rs751037529 | 395419 | Autism_spectrum_disorder\|not_provided\|Autosomal_recessive_juvenile_Parkinson_disease_2 | MONDO:MONDO:0005258,MeSH:D0 | criteria_provided,_single_submitter | Pathogenic | 0 | 0.912 | D |
| chr6 | 161785839 | 161785839 | A | T | exonic | PRKN | . | stopgain | PRKN:NM_013988:exon4:c.T357A:p.C119X,<br>PRKN:NM_013987:exon6:c.T720A:p.C240X,<br>PRKN:NM_004562:exon7:c.T804A:p.C268X | rs377554392 | 793178 | not_provided | MedGen:C3661900 | criteria_provided,_multiple_submitters,_no_conflicts | Pathogenic | . | . | . |
| chr6 | 162443314 | 162443314 | A | T | exonic | PRKN | . | nonsynonymous SNV | PRKN:NM_004562:exon2:c.T167A:p.V56E,<br>PRKN:NM_013987:exon2:c.T167A:p.V56E,<br>PRKN:NM_013988:exon2:c.T167A:p.V56E | rs137853059 | 22086 | not_provided\|Autosomal_recessive_juvenile_Parkinson_disease_2 | MedGen:CN517202\|MONDO:MOND | criteria_provided,_multiple_submitters,_no_conflicts | Pathogenic | 0.001 | 0.784 | D |
| chr6 | 162443356 | 162443356 | C | G | exonic | PRKN | . | nonsynonymous SNV | PRKN:NM_004562:exon2:c.G125C:p.R42P,<br>PRKN:NM_013987:exon2:c.G125C:p.R42P,<br>PRKN:NM_013988:exon2:c.G125C:p.R42P | rs368134308 | 634774 | not_provided | MedGen:C3661900 | criteria_provided,_single_submitter | Pathogenic | 0.011 | 0.564 | D |

| Polyphen2 HDIV score | Polyphen2 HDIV rankscore | Polyphen2 HDIV pred | Polyphen2 HVAR score | Polyphen2 HVAR rankscore | Polyphen2 HVAR pred | LRT score | LRT converted rankscore | LRT pred | MutationTaster score | MutationTaster converted rankscore | MutationTaster pred | MutationAssessor score | MutationAssessor score rankscore | MutationAssessor pred | FATHMM score | FATHMM converted rankscore | FATHMM pred | PROVEAN score | PROVEAN converted rankscore | PROVEAN pred | VEST3 score | VEST3 rankscore | MetaSVM score | MetaSVM rankscore | MetaSVM pred | MetaLR score | MetaLR rankscore | MetaLR pred | M-CAP score | M-CAP rankscore | M-CAP pred | CADD raw | CADD raw rankscore | CADD phred | DANN score | DANN rankscore | fathmm-MKL coding score | fathmm-MKL coding rankscore | fathmm-MKL coding pred |
|---|---|---|---|---|---|---|---|---|---|---|---|---|---|---|---|---|---|---|---|---|---|---|---|---|---|---|---|---|---|---|---|---|---|---|---|---|---|---|---|
| 1 | 0.899 | D | 0.994 | 0.875 | D | 0.001 | 0.424 | D | 1 | 0.473 | D | 2.455 | 0.715 | M | -1.38 | 0.803 | T | -4.5 | 0.781 | D | 0.924 | 0.916 | 0.245 | 0.867 | D | 0.629 | 0.87 | D | 0.19 | 0.862 | D | 5.652 | 0.77 | 26.7 | 0.999 | 0.994 | 0.776 | 0.379 | D |
| . | . | . | . | . | . | . | . | . | . | . | . | . | . | . | . | . | . | . | . | . | . | . | . | . | . | . | . | . | . | . | . | . | . | . | . | . | . | . | . |
| 1 | 0.899 | D | 0.999 | 0.916 | D | 0 | 0.843 | D | 1 | 0.81 | D | 3.54 | 0.933 | H | -5 | 0.985 | D | -7.91 | 0.962 | D | 0.988 | 0.992 | 1.091 | 0.993 | D | 0.973 | 0.991 | D | 0.542 | 0.958 | D | 7.949 | 0.956 | 35 | 0.999 | 0.998 | 0.997 | 0.986 | D |
| 1 | 0.899 | D | 1 | 0.971 | D | 0 | 0.843 | D | 1 | 0.81 | A | 3.545 | 0.934 | H | -5.3 | 0.99 | D | -5.93 | 0.89 | D | 0.987 | 0.991 | 1.065 | 0.985 | D | 0.979 | 0.993 | D | 0.546 | 0.958 | D | 7.354 | 0.949 | 34 | 0.999 | 0.944 | 0.994 | 0.96 | D |
| 1 | 0.899 | D | 0.998 | 0.875 | D | 0 | 0.843 | D | 1 | 0.588 | D | 2.65 | 0.778 | M | -4.51 | 0.977 | D | -5.83 | 0.883 | D | 0.939 | 0.933 | 1.091 | 0.993 | D | 0.954 | 0.985 | D | 0.327 | 0.917 | D | 7.163 | 0.944 | 34 | 0.999 | 0.989 | 0.871 | 0.463 | D |
| 1 | 0.899 | D | 0.998 | 0.875 | D | 0 | 0.843 | D | 1 | 0.81 | D | 2.845 | 0.829 | M | -2.27 | 0.874 | D | -6.39 | 0.911 | D | 0.921 | 0.913 | 0.969 | 0.967 | D | 0.876 | 0.959 | D | 0.328 | 0.917 | D | 7.906 | 0.956 | 35 | 0.999 | 0.989 | 0.985 | 0.838 | D |
| . | . | . | . | . | . | . | . | . | 1 | 0.81 | D | . | . | . | . | . | . | . | . | . | . | . | . | . | . | . | . | . | . | . | . | 5.333 | 0.722 | 25.8 | 0.996 | 0.717 | 0.918 | 0.541 | D |
| 1 | 0.899 | D | 0.996 | 0.832 | D | 0 | 0.843 | D | 1 | 0.81 | A | 1.95 | 0.526 | M | -3.1 | 0.927 | D | -3.27 | 0.655 | D | 0.91 | 0.9 | 0.779 | 0.942 | D | 0.832 | 0.944 | D | 0.215 | 0.875 | D | 6.527 | 0.899 | 31 | 0.999 | 0.996 | 0.99 | 0.902 | D |
| 1 | 0.899 | D | 0.997 | 0.85 | D | 0 | 0.843 | D | 1 | 0.81 | D | 3.555 | 0.935 | H | -3.39 | 0.942 | D | -6.42 | 0.912 | D | 0.972 | 0.973 | 1.039 | 0.979 | D | 0.925 | 0.975 | D | 0.226 | 0.881 | D | 7.881 | 0.956 | 35 | 0.999 | 0.995 | 0.988 | 0.872 | D |
| 0.999 | 0.764 | D | 0.932 | 0.65 | D | 0.003 | 0.361 | N | 1 | 0.588 | A | 2.705 | 0.794 | M | -5.84 | 0.994 | D | -5.28 | 0.843 | D | 0.903 | 0.893 | 1.058 | 0.983 | D | 0.968 | 0.99 | D | 0.917 | 0.994 | D | 6.394 | 0.883 | 29.6 | 0.999 | 0.966 | 0.829 | 0.418 | D |
| 0.999 | 0.899 | D | 0.97 | 0.764 | D | 0 | 0.457 | D | 0.968 | 0.423 | D | 2.67 | 0.784 | M | -5.72 | 0.993 | D | -2.13 | 0.5 | N | 0.569 | 0.778 | 1.088 | 0.992 | D | 0.968 | 0.99 | D | 0.886 | 0.991 | D | 5.422 | 0.735 | 26 | 0.999 | 0.982 | 0.7 | 0.341 | D |
| 1 | 0.899 | D | 0.995 | 0.832 | D | 0 | 0.629 | D | 1 | 0.513 | A | 3.29 | 0.905 | M | -6.24 | 0.996 | D | -3.57 | 0.705 | D | 0.896 | 0.885 | 1.063 | 0.984 | D | 0.984 | 0.995 | D | 0.89 | 0.992 | D | 6.932 | 0.933 | 33 | 0.999 | 0.999 | 0.964 | 0.685 | D |
| 0.001 | 0.147 | B | 0.001 | 0.154 | B | 0.006 | 0.321 | N | 1 | 0.588 | D | 1.215 | 0.307 | L | -5.8 | 0.994 | D | -3.81 | 0.718 | D | 0.915 | 0.906 | 0.508 | 0.907 | D | 0.816 | 0.938 | D | 0.422 | 0.938 | D | -0.512 | 0.055 | 0.206 | 0.639 | 0.071 | 0.913 | 0.53 | D |
| 0.001 | 0.102 | B | 0.019 | 0.176 | B | 0 | 0.537 | N | 1 | 0.479 | A | 1.56 | 0.396 | L | -5.71 | 0.993 | D | -3.05 | 0.664 | D | 0.366 | 0.427 | 0.425 | 0.895 | D | 0.843 | 0.948 | D | 0.241 | 0.887 | D | -1.034 | 0.031 | 0.013 | 0.46 | 0.036 | 0.816 | 0.407 | D |
| 0.969 | 0.764 | D | 0.874 | 0.668 | P | 0 | 0.629 | D | 1 | 0.588 | D | 3.67 | 0.945 | H | -7.03 | 0.998 | D | -6.84 | 0.93 | D | 0.911 | 0.906 | 0.979 | 0.968 | D | 0.993 | 0.998 | D | 0.807 | 0.985 | D | 6.19 | 0.854 | 28.6 | 0.998 | 0.894 | 0.966 | 0.697 | D |
| 1 | 0.899 | D | 1 | 0.971 | D | 0 | 0.457 | N | 1 | 0.81 | D | 2.91 | 0.844 | M | -6.3 | 0.996 | D | -6.88 | 0.932 | D | 0.885 | 0.873 | 1.085 | 0.99 | D | 0.975 | 0.992 | D | 0.885 | 0.991 | D | 4.813 | 0.646 | 24.8 | 0.997 | 0.774 | 0.873 | 0.466 | D |
| 1 | 0.899 | D | 1 | 0.971 | D | 0 | 0.843 | D | 1 | 0.537 | D | 4.445 | 0.988 | H | -2.62 | 1 | D | -7.63 | 0.964 | D | 0.984 | 0.987 | 0.913 | 0.959 | D | 0.998 | 1 | D | 0.869 | 0.99 | D | 7.438 | 0.951 | 34 | 0.999 | 0.979 | 0.993 | 0.936 | D |
| . | . | . | . | . | . | 0.491 | 0.121 | N | 1 | 0.81 | A | . | . | . | . | . | . | . | . | . | . | . | . | . | . | . | . | . | . | . | . | 9.496 | 0.963 | 35 | 0.996 | 0.75 | 0.089 | 0.146 | N |
| . | . | . | . | . | . | 0 | 0.843 | D | 1 | 0.81 | A | . | . | . | . | . | . | . | . | . | . | . | . | . | . | . | . | . | . | . | . | 11.471 | 0.975 | 37 | 0.998 | 0.85 | 0.998 | 0.993 | D |
| . | . | . | . | . | . | 0 | 0.843 | D | 1 | 0.81 | A | . | . | . | . | . | . | . | . | . | . | . | . | . | . | . | . | . | . | . | . | 10.346 | 0.967 | 36 | 0.997 | 0.833 | 0.943 | 0.606 | D |
| . | . | . | . | . | . | 0 | 0.481 | D | 1 | 0.81 | A | . | . | . | . | . | . | . | . | . | . | . | . | . | . | . | . | . | . | . | . | 10.303 | 0.967 | 36 | 0.993 | 0.599 | 0.777 | 0.379 | D |
| 1 | 0.899 | D | 0.999 | 0.971 | D | 0 | 0.843 | D | 1 | 0.81 | A | 3.84 | 0.958 | H | -1.18 | 0.783 | T | -6.67 | 0.923 | D | 0.94 | 0.934 | 0.775 | 0.941 | D | 0.739 | 0.911 | D | 0.299 | 0.909 | D | 5.08 | 0.684 | 25.3 | 0.998 | 0.914 | 0.97 | 0.718 | D |
| . | . | . | . | . | . | 0.145 | 0.181 | N | 1 | 0.81 | A | . | . | . | . | . | . | . | . | . | . | . | . | . | . | . | . | . | . | . | . | 12.745 | 0.986 | 40 | 0.997 | 0.822 | 0.79 | 0.388 | D |
| . | . | . | . | . | . | 0 | 0.843 | D | 1 | 0.81 | D | . | . | . | . | . | . | . | . | . | . | . | . | . | . | . | . | . | . | . | . | 15.572 | 0.999 | 51 | 0.998 | 0.848 | 0.931 | 0.571 | D |
| 0.89 | 0.622 | P | 0.716 | 0.608 | P | 0 | 0.629 | D | 1 | 0.81 | D | 3.005 | 0.862 | M | 0.8 | 0.488 | T | -5.58 | 0.864 | D | 0.889 | 0.877 | -0.396 | 0.722 | T | 0.291 | 0.663 | T | 0.039 | 0.586 | D | 5.927 | 0.814 | 27.6 | 0.998 | 0.882 | 0.853 | 0.441 | D |
| . | . | . | . | . | . | . | . | . | 1 | 0.81 | A | . | . | . | . | . | . | . | . | . | . | . | . | . | . | . | . | . | . | . | . | 10.948 | 0.971 | 37 | 0.994 | 0.629 | 0.615 | 0.312 | D |
| . | . | . | . | . | . | 0 | 0.559 | D | 1 | 0.81 | A | . | . | . | . | . | . | . | . | . | . | . | . | . | . | . | . | . | . | . | . | 14.038 | 0.995 | 44 | 0.998 | 0.899 | 0.992 | 0.933 | D |
| 1 | 0.899 | D | 0.995 | 0.818 | D | 0 | 0.559 | D | 1 | 0.588 | D | 2.195 | 0.62 | M | -0.3 | 0.679 | T | -4.43 | 0.775 | D | 0.879 | 0.867 | 0.147 | 0.85 | D | 0.536 | 0.829 | D | 0.228 | 0.882 | D | 7.28 | 0.948 | 34 | 0.996 | 0.728 | 0.989 | 0.876 | D |
| . | . | . | . | . | . | 0.795 | 0.094 | U | 1 | 0.81 | A | . | . | . | . | . | . | . | . | . | . | . | . | . | . | . | . | . | . | . | . | 8.185 | 0.957 | 35 | 0.998 | 0.879 | 0.113 | 0.164 | N |
| . | . | . | . | . | . | . | . | . | . | . | . | . | . | . | . | . | . | . | . | . | . | . | . | . | . | . | . | . | . | . | . | . | . | . | . | . | . | . | . |
| 1 | 0.899 | D | 1 | 0.971 | D | 0 | 0.843 | D | 1 | 0.499 | D | 2.44 | 0.71 | M | -7 | 0.998 | D | -6.9 | 0.944 | D | 0.966 | 0.969 | 0.976 | 0.968 | D | 0.992 | 0.998 | D | 0.494 | 0.951 | D | 6.786 | 0.924 | 32 | 0.999 | 0.998 | 0.934 | 0.58 | D |
| . | . | . | . | . | . | 0 | 0.843 | D | 1 | 0.81 | A | . | . | . | . | . | . | . | . | . | . | . | . | . | . | . | . | . | . | . | . | 9.852 | 0.964 | 36 | 0.986 | 0.429 | 0.312 | 0.241 | N |
| 0.987 | 0.715 | D | 0.88 | 0.728 | P | 0.002 | 0.369 | N | 0.977 | 0.81 | A | 2.33 | 0.67 | M | -4.02 | 0.964 | D | -4.3 | 0.789 | D | 0.94 | 0.934 | 1.06 | 0.983 | D | 0.918 | 0.973 | D | 0.402 | 0.934 | D | 6.194 | 0.855 | 28.6 | 0.988 | 0.461 | 0.919 | 0.542 | D |
| 0.998 | 0.715 | D | 0.96 | 0.719 | D | 0 | 0.843 | D | 0.988 | 0.81 | D | 3.335 | 0.911 | M | -1.19 | 0.784 | T | -3.86 | 0.752 | D | 0.968 | 0.968 | 0.502 | 0.906 | D | 0.674 | 0.887 | D | 0.124 | 0.806 | D | 7.095 | 0.941 | 33 | 0.998 | 0.868 | 0.808 | 0.401 | D |

| Eigen coding or noncoding | Eigen-raw | Eigen-PC raw | GenoCanyon score | GenoCanyonscore rankscore | integrated fitCons score | integrated fitCons score rankscore | integrated confidence value | GERP++ RS | GERP++ RS rankscore | phyloP100way vertebrate | phyloP100way vertebrate rankscore | phyloP20way mammalian | phyloP20way mammalian rankscore | phastCons100way vertebrate | phastCons100way vertebrate rankscore | phastCons20way mammalian | phastCons20way mammalia rankscore | SiPhy 29way logOdds | SiPhy 29way logOdds rankscore | Interpro domain |
|---|---|---|---|---|---|---|---|---|---|---|---|---|---|---|---|---|---|---|---|---|
| c | 0.361 | 0.259 | 0.999 | 0.396 | 0.554 | 0.246 | 0 | 4.74 | 0.596 | 3.725 | 0.544 | 0.852 | 0.362 | 0.998 | 0.411 | 0.885 | 0.372 | 15.915 | 0.792 | Mitochondrial Rho-like\|P-loop containing nucleoside triphosphate hydrolase\|Roc domain\|Small GTP-binding protein domain |
| . | . | . | . | . | . | . | . | . | . | . | . | . | . | . | . | . | . | . | . | . |
| c | 1.009 | 0.949 | 1 | 0.983 | 0.672 | 0.522 | 0 | 5.5 | 0.813 | 9.508 | 0.97 | 1.048 | 0.713 | 1 | 0.715 | 0.994 | 0.587 | 19.399 | 0.946 | DNA-directed DNA polymerase, family A, palm domain |
| c | 0.811 | 0.719 | 1 | 0.747 | 0.672 | 0.522 | 0 | 4.56 | 0.555 | 7.498 | 0.802 | 0.935 | 0.49 | 1 | 0.715 | 0.995 | 0.604 | 15.416 | 0.746 | DNA-directed DNA polymerase, family A, palm domain |
| c | 0.589 | 0.504 | 1 | 0.747 | 0.707 | 0.73 | 0 | 4.05 | 0.462 | 2.763 | 0.47 | 1.048 | 0.713 | 1 | 0.715 | 1 | 0.888 | 13.225 | 0.592 | . |
| c | 0.691 | 0.674 | 1 | 0.983 | 0.707 | 0.73 | 0 | 4.99 | 0.658 | 8.04 | 0.891 | 1.048 | 0.713 | 1 | 0.715 | 1 | 0.888 | 18.267 | 0.899 | . |
| c | 0.997 | 0.824 | 1 | 0.983 | 0.284 | 0.042 | 0 | 4.99 | 0.658 | 7.271 | 0.777 | 0.892 | 0.403 | 1 | 0.715 | 0.888 | 0.373 | 18.463 | 0.907 | . |
| c | 0.662 | 0.641 | 1 | 0.983 | 0.707 | 0.73 | 0 | 5 | 0.661 | 7.468 | 0.798 | 0.892 | 0.403 | 1 | 0.715 | 0.98 | 0.49 | 18.482 | 0.907 | . |
| c | 1.071 | 1.022 | 1 | 0.983 | 0.707 | 0.73 | 0 | 5.84 | 0.934 | 10.003 | 0.997 | 1.048 | 0.713 | 1 | 0.715 | 1 | 0.888 | 20.135 | 0.98 | Ribonuclease H-like domain |
| c | 0.306 | 0.17 | 0.273 | 0.189 | 0.706 | 0.609 | 0 | 2.11 | 0.262 | 1.048 | 0.298 | 0.803 | 0.325 | 1 | 0.715 | 0.722 | 0.315 | 9.221 | 0.364 | Glycoside hydrolase superfamily\|Glycoside hydrolase, catalytic domain;Glycosyl hydrolase, family 13, all-beta |
| c | 0.4 | 0.293 | 0.988 | 0.314 | 0.707 | 0.73 | 0 | 3.67 | 0.411 | 2.224 | 0.423 | 0.914 | 0.427 | 0.808 | 0.296 | 0.988 | 0.529 | 11.005 | 0.467 | Glycoside hydrolase superfamily\|Glycoside hydrolase, catalytic domain |
| c | 0.638 | 0.512 | 1 | 0.489 | 0.706 | 0.609 | 0 | 3.51 | 0.391 | 5.235 | 0.65 | 0.818 | 0.335 | 1 | 0.715 | 0.999 | 0.75 | 10.668 | 0.447 | Glycoside hydrolase superfamily\|Glycoside hydrolase, catalytic domain |
| c | -0.862 | -0.747 | 0.985 | 0.308 | 0.706 | 0.609 | 0 | 2.5 | 0.292 | 4.227 | 0.582 | 1.053 | 0.755 | 0.995 | 0.385 | 0.712 | 0.312 | 7.355 | 0.257 | Glycoside hydrolase superfamily\|Glycoside hydrolase, catalytic domain |
| c | -0.761 | -0.71 | 0.859 | 0.252 | 0.706 | 0.609 | 0 | 1.25 | 0.204 | 3.36 | 0.518 | 0.935 | 0.49 | 1 | 0.715 | 0.804 | 0.337 | 7.407 | 0.26 | Glycoside hydrolase superfamily\|Glycoside hydrolase, catalytic domain |
| c | 0.57 | 0.412 | 0.282 | 0.19 | 0.706 | 0.609 | 0 | 3.55 | 0.396 | 4.353 | 0.589 | 0.051 | 0.162 | 1 | 0.715 | 0.99 | 0.544 | 10.764 | 0.453 | Glycoside hydrolase superfamily\|Glycoside hydrolase, catalytic domain;Glycoside hydrolase, catalytic domain |
| c | 0.001 | -0.215 | 1 | 0.411 | 0.732 | 0.924 | 0 | 1.33 | 0.209 | 2.842 | 0.477 | 0.048 | 0.16 | 1 | 0.715 | 0.025 | 0.141 | 3.812 | 0.083 | Glycosyl hydrolase, family 13, all-beta |
| c | 0.818 | 0.661 | 1 | 0.5 | 0.713 | 0.817 | 0 | 3.46 | 0.386 | 7.512 | 0.805 | 0.855 | 0.374 | 1 | 0.715 | 0.997 | 0.653 | 9.609 | 0.386 | HAD-like domain |
| c | 0.249 | -0.059 | 0.977 | 0.297 | 0.706 | 0.609 | 0 | 2.53 | 0.295 | -0.032 | 0.121 | 0.953 | 0.551 | 0 | 0.063 | 0.109 | 0.192 | 8.173 | 0.302 | . |
| c | 1.031 | 0.869 | 1 | 0.747 | 0.706 | 0.609 | 0 | 5.73 | 0.897 | 9.905 | 0.986 | 0.994 | 0.605 | 1 | 0.715 | 0.988 | 0.529 | 18.453 | 0.906 | P-type ATPase, cytoplasmic domain N |
| c | 0.556 | 0.36 | 0.371 | 0.199 | 0.706 | 0.609 | 0 | 4.27 | 0.498 | 1.185 | 0.315 | 0.994 | 0.605 | 0.922 | 0.318 | 0.984 | 0.507 | 11.753 | 0.509 | P-type ATPase, A domain |
| c | 0.704 | 0.546 | 0.983 | 0.305 | 0.707 | 0.73 | 0 | 3.44 | 0.383 | 3.246 | 0.509 | 0.824 | 0.337 | 1 | 0.715 | 0.985 | 0.512 | 9.358 | 0.372 | . |
| c | 0.734 | 0.578 | 0.999 | 0.374 | 0.707 | 0.73 | 0 | 4.9 | 0.635 | 6.966 | 0.757 | 1.058 | 0.762 | 1 | 0.715 | 0.043 | 0.159 | 10.397 | 0.432 | Protein kinase domain\|Protein kinase-like domain |
| c | 0.735 | 0.59 | 0.95 | 0.278 | 0.672 | 0.522 | 0 | 5.13 | 0.696 | 2.193 | 0.42 | 0.935 | 0.49 | 0.861 | 0.304 | 0.955 | 0.433 | 13.364 | 0.599 | Protein kinase domain\|Protein kinase-like domain |
| c | 0.894 | 0.8 | 0.851 | 0.25 | 0.672 | 0.522 | 0 | 6.17 | 0.997 | 2.897 | 0.481 | 0.935 | 0.49 | 1 | 0.715 | 1 | 0.888 | 16.38 | 0.832 | Protein kinase domain\|Protein kinase-like domain |
| c | 0.578 | 0.509 | 1 | 0.747 | 0.778 | 0.996 | 0 | 3.87 | 0.437 | 4.33 | 0.588 | 0.847 | 0.346 | 0.999 | 0.424 | 0.998 | 0.697 | 11.896 | 0.518 | . |
| c | 0.866 | 0.681 | 1 | 0.747 | 0.493 | 0.174 | 0 | 3.58 | 0.4 | 2.509 | 0.449 | 0.934 | 0.45 | 0.922 | 0.318 | 1 | 0.888 | 12.369 | 0.544 | . |
| c | 0.867 | 0.724 | 1 | 0.747 | 0.706 | 0.609 | 0 | 4.53 | 0.548 | 7.506 | 0.804 | 0.953 | 0.551 | 1 | 0.715 | 0.998 | 0.697 | 17.634 | 0.88 | Acyl transferase/acyl hydrolase/lysophospholipase\|Patatin/Phospholipase A2-related |
| c | 0.798 | 0.781 | 1 | 0.747 | 0.707 | 0.73 | 0 | 5.49 | 0.809 | 7.358 | 0.786 | 0.935 | 0.49 | 1 | 0.715 | 0.983 | 0.502 | 19.374 | 0.945 | Ankyrin repeat-containing domain |
| c | 0.355 | 0.066 | 1 | 0.48 | 0.706 | 0.609 | 0 | 4.63 | 0.57 | 1.884 | 0.391 | 0.953 | 0.551 | 0.185 | 0.239 | 0.29 | 0.235 | 14.39 | 0.664 | . |
| . | . | . | . | . | . | . | . | . | . | . | . | . | . | . | . | . | . | . | . | . |
| c | 0.766 | 0.697 | 1 | 0.431 | 0.554 | 0.246 | 0 | 5.75 | 0.904 | 6.982 | 0.758 | 0.935 | 0.49 | 1 | 0.715 | 0.923 | 0.397 | 17.711 | 0.882 | . |
| c | -0.131 | -0.413 | 0 | 0.065 | 0.554 | 0.246 | 0 | -6.67 | 0.016 | -0.202 | 0.094 | 0.234 | 0.26 | 0.533 | 0.27 | 0.999 | 0.75 | 15.684 | 0.771 | . |
| c | 0.732 | 0.732 | 1 | 0.489 | 0.516 | 0.203 | 0 | 5.63 | 0.861 | 7.205 | 0.772 | 1.199 | 0.96 | 1 | 0.715 | 0.995 | 0.604 | 16.131 | 0.812 | Ubiquitin domain\|Ubiquitin-related domain |
| c | 0.785 | 0.709 | 0.994 | 0.336 | 0.554 | 0.246 | 0 | 4.74 | 0.596 | 0.748 | 0.258 | 0.935 | 0.49 | 0.996 | 0.391 | 1 | 0.888 | 15.471 | 0.751 | Ubiquitin domain\|Ubiquitin-related domain |
