## Supplementary Table 3 for "Parkinson’s Disease Pathogenic Variants: Cross-Ancestry Analysis and Microarray Data Validation"

**Typed Pathogenic Variant Cluster Plot Classification**

**Table 3A**

| SNP | rsID | Gene | Classification | Number of NC | GP2 r7 Missing | GP2 r7 MAF |
|---|---|---|---|---|---|---|
| chr1:16988455:C:T | rs144701072 | ATP13A2 | Bad | 20 | 0.017351 | 0.001302 |
| chr1:16989961:G:A | rs866035312 | ATP13A2 | Bad | 11 | 0.017162 | 0.00416 |
| chr1:20645640:T:C | rs28940285 | PINK1 | Bad | 10 | 0.003482 | 0.000517 |
| chr1:20649109:C:T | rs45539432 | PINK1 | Good | 0 | 0.006364 | 0.001309 |
| chr1:20649217:C:T | rs34208370 | PINK1 | Bad | 47 | 0.017892 | 0.003796 |
| chr1:155235196:G:A | rs80356771 | GBA | Good | 1 | 0.002296 | 0.00132 |
| chr1:155237453:C:T | rs78973108 | GBA | Good | 2 | 0.003762 | 0.001097 |
| chr1:155238206:A:C | rs381427 | GBA | Bad | 23 | 0.008074 | 0.000208 |
| chr1:155238215:T:C | rs364897 | GBA | Good | 0 | 0.001512 | 0.000275 |
| chr6:161785793:C:G | rs751037529 | PRKN | Good | 0 | 0.002405 | 0.000103 |
| chr6:161785839:A:T | rs377554392 | PRKN | Good | 1 | 0.001941 | 0.000103 |
| chr6:162443314:A:T | rs137853059 | PRKN | Good | 0 | 0.00219 | 0.00025 |
| chr6:161350214:G:C | rs765860776 | PRKN | Good | 4 | 0.003882 | 0.000759 |
| chr6:162443356:C:G | rs368134308 | PRKN | Good | 1 | 0.002835 | 0.00019 |
| chr12:40310434:C:T | rs33939927 | LRRK2 | Good | 4 | 0.003917 | 0.001251 |
| chr15:89321780:G:A | rs144500145 | POLG | Bad | 66 | 0.017689 | 0.004101 |
| chr15:89321792:C:T | rs113994098 | POLG | Bad | 45 | 0.016234 | 0.002766 |
| chr15:89325610:G:A | rs139717885 | POLG | Good | 1 | 0.004681 | 0.000889 |
| chr15:89325679:G:A | rs774474723 | POLG | Medium | 3 | 0.004854 | 0.000691 |
| chr15:89327166:C:T | rs771623994 | POLG | Medium | 8 | 0.006459 | 0.001431 |
| chr15:89327201:C:T | rs113994095 | POLG | Bad | 13 | 0.008018 | 0.002619 |
| chr15:89329041:G:A | rs886041592 | POLG | Medium | 4 | 0.009293 | 0.002384 |
| chr22:38115658:G:A | rs587784339 | PLA2G6 | Bad | 98 | 0.02223 | 0.001729 |
| chr22:38132917:C:A | rs199935023 | PLA2G6 | Good | 4 | 0.005746 | 0.000307 |
| chr22:38169318:G:A | rs200075782 | PLA2G6 | Medium | 6 | 0.006914 | 0.002151 |

**Table 3B**

| Classification | N | Mean NC | Min NC | Max NC | Avg. Missingne | Avg. MAF |
|---|---|---|---|---|---|---|
| Good | 12 | 1.5 | 0 | 4 | 0.003461 | 0.000654 |
| Medium | 4 | 5.25 | 3 | 8 | 0.00688 | 0.001664 |
| Bad | 9 | 37 | 10 | 98 | 0.014236 | 0.002355 |
