## Supplementary Table 4 for "Parkinson’s Disease Pathogenic Variants: Cross-Ancestry Analysis and Microarray Data Validation"

| **gnomAD (v4.1.0) pathogenic variant information for Parkinson's Disease genes** | | | | | | | | | | | | |
|---|---|---|---|---|---|---|---|---|---|---|---|---|
| gene | gnomAD_ID | Chromosome | Position | rsIDs | Reference | Alternate | ClinVar_ClnSignificance | ClinVar_varID | allele_count | allele_number | allele_freq | in_gp2 |
| ATP13A2 | 1-16986881-CAG-C | 1 | 16986881 | rs1570759415 | CAG | C | Pathogenic | 660925 | 1 | 1614048 | 6.20E-07 | FALSE |
| ATP13A2 | 1-16986885-GAGA-G | 1 | 16986885 | rs1057519290 | GAGA | G | Pathogenic | 374887 | 1 | 1614104 | 6.20E-07 | FALSE |
| ATP13A2 | 1-16986886-A-AG | 1 | 16986886 | rs747617559 | A | AG | Pathogenic | 1968613 | 2 | 1614040 | 1.24E-06 | FALSE |
| ATP13A2 | 1-16988455-C-T | 1 | 16988455 | rs144701072 | C | T | Pathogenic | 66099 | 15 | 1613774 | 9.29E-06 | TRUE |
| ATP13A2 | 1-16989739-A-C | 1 | 16989739 | rs587777053 | A | C | Pathogenic | 66098 | 1 | 1614078 | 6.20E-07 | FALSE |
| ATP13A2 | 1-16989961-G-A | 1 | 16989961 | rs866035312 | G | A | Pathogenic | 502116 | 9 | 1607028 | 5.60E-06 | TRUE |
| ATP13A2 | 1-16992022-G-A | 1 | 16992022 | rs774115028 | G | A | Pathogenic | 1996186 | 1 | 1612960 | 6.20E-07 | FALSE |
| ATP13A2 | 1-16992315-CT-C | 1 | 16992315 | rs2076962370 | CT | C | Pathogenic | 2923611 | 2 | 1612322 | 1.24E-06 | FALSE |
| ATP13A2 | 1-16992345-G-A | 1 | 16992345 | rs773246271 | G | A | Pathogenic | 465252 | 46 | 1613016 | 2.85E-05 | TRUE |
| ATP13A2 | 1-16992506-C-A | 1 | 16992506 | | C | A | Pathogenic | 1999337 | 1 | 1614160 | 6.20E-07 | FALSE |
| ATP13A2 | 1-16996059-G-A | 1 | 16996059 | rs1303653650 | G | A | Pathogenic | 520750 | 20 | 1613950 | 1.24E-05 | TRUE |
| ATP13A2 | 1-16996262-G-A | 1 | 16996262 | rs1057519293 | G | A | Pathogenic | 374890 | 15 | 1614168 | 9.29E-06 | FALSE |
| ATP13A2 | 1-16997101-GC-G | 1 | 16997101 | rs1377055875 | GC | G | Pathogenic | 1385899 | 3 | 1613740 | 1.86E-06 | FALSE |
| ATP13A2 | 1-16997112-G-GTC | 1 | 16997112 | rs762033589 | G | GTC | Pathogenic | 30833 | 1 | 1613832 | 6.20E-07 | FALSE |
| ATP13A2 | 1-17000466-C-T | 1 | 17000466 | rs1334843918 | C | T | Pathogenic | 2046647 | 1 | 1613948 | 6.20E-07 | FALSE |
| ATP13A2 | 1-17002312-G-A | 1 | 17002312 | rs1483668823 | G | A | Pathogenic | 1442231 | 1 | 1613710 | 6.20E-07 | FALSE |
| ATP13A2 | 1-17002326-TG-T | 1 | 17002326 | rs758150853 | TG | T | Pathogenic | 1751269 | 7 | 1613436 | 4.34E-06 | FALSE |
| ATP13A2 | 1-17004817-C-A | 1 | 17004817 | rs756152157 | C | A | Pathogenic | 3336424 | 5 | 1613772 | 3.10E-06 | FALSE |
| ATP13A2 | 1-17005444-AC-A | 1 | 17005444 | rs1389678247 | AC | A | Pathogenic | 2931003 | 4 | 1613712 | 2.48E-06 | FALSE |
| ATP13A2 | 1-17005449-C-T | 1 | 17005449 | | C | T | Pathogenic | 2419737 | 1 | 1614170 | 6.20E-07 | FALSE |
| ATP13A2 | 1-17005450-C-T | 1 | 17005450 | rs373607247 | C | T | Pathogenic | 432661 | 14 | 1614062 | 8.67E-06 | TRUE |
| FBXO7 | 22-32475003-A-C | 22 | 32475003 | rs753392528 | A | C | Pathogenic | 2887235 | 18 | 1536094 | 1.17E-05 | FALSE |
| FBXO7 | 22-32475004-T-G | 22 | 32475004 | rs945794813 | T | G | Pathogenic | 2916108 | 10 | 1536252 | 6.51E-06 | FALSE |
| FBXO7 | 22-32475004-T-A | 22 | 32475004 | | T | A | Pathogenic | 2960687 | 1 | 1536252 | 6.51E-07 | FALSE |
| FBXO7 | 22-32475067-C-T | 22 | 32475067 | rs121918305 | C | T | Pathogenic | 4811 | 7 | 1546034 | 4.53E-06 | FALSE |
| FBXO7 | 22-32478991-C-T | 22 | 32478991 | rs1370252127 | C | T | Pathogenic | 2661902 | 17 | 1613960 | 1.05E-05 | FALSE |
| FBXO7 | 22-32479008-GA-G | 22 | 32479008 | rs1228608709 | GA | G | Pathogenic | 488517 | 4 | 1614138 | 2.48E-06 | FALSE |
| FBXO7 | 22-32479126-TCC-T | 22 | 32479126 | rs750398883 | TCC | T | Pathogenic | 2150213 | 21 | 1614180 | 1.30E-05 | FALSE |
| FBXO7 | 22-32479233-AG-A | 22 | 32479233 | rs2057448454 | AG | A | Pathogenic | 2884317 | 3 | 1614158 | 1.86E-06 | FALSE |
| FBXO7 | 22-32483976-C-G | 22 | 32483976 | rs779737534 | C | G | Pathogenic | 2751184 | 1 | 1614208 | 6.19E-07 | FALSE |
| FBXO7 | 22-32484021-A-AT | 22 | 32484021 | | A | AT | Pathogenic | 2757154 | 2 | 1614194 | 1.24E-06 | FALSE |
| FBXO7 | 22-32484052-CTG-C | 22 | 32484052 | rs778770873 | CTG | C | Pathogenic | 2699868 | 1 | 1614176 | 6.20E-07 | FALSE |
| FBXO7 | 22-32485131-C-T | 22 | 32485131 | rs2057490468 | C | T | Pathogenic | 2733394 | 14 | 1614194 | 8.67E-06 | FALSE |
| FBXO7 | 22-32487774-AGT-A | 22 | 32487774 | rs1157030138 | AGT | A | Pathogenic | 2900056 | 3 | 1609598 | 1.86E-06 | FALSE |
| FBXO7 | 22-32491107-A-AT | 22 | 32491107 | rs2057531939 | A | AT | Pathogenic | 2960440 | 2 | 1613280 | 1.24E-06 | FALSE |
| FBXO7 | 22-32493278-C-T | 22 | 32493278 | rs78099169 | C | T | Pathogenic | 2911981 | 8 | 1613536 | 4.96E-06 | FALSE |
| FBXO7 | 22-32498174-G-T | 22 | 32498174 | rs1342038737 | G | T | Pathogenic | 1455741 | 14 | 1614112 | 8.67E-06 | FALSE |
| FBXO7 | 22-32498177-TC-T | 22 | 32498177 | rs2057587984 | TC | T | Pathogenic | 2715290 | 18 | 1614048 | 1.12E-05 | FALSE |
| FBXO7 | 22-32498410-TC-T | 22 | 32498410 | rs1344213916 | TC | T | Pathogenic | 2977139 | 3 | 1614172 | 1.86E-06 | FALSE |
| FBXO7 | 22-32498453-C-T | 22 | 32498453 | rs121918304 | C | T | Pathogenic | 4809 | 16 | 1614170 | 9.91E-06 | FALSE |
| GBA1 | 1-155235196-G-A | 1 | 155235196 | rs80356771 | G | A | Pathogenic | 4295 | 370 | 1612378 | 0.00022947472 | TRUE |
| GBA1 | 1-155235680-C-T | 1 | 155235680 | rs1671699033 | C | T | Pathogenic | 1322984 | 7 | 1598222 | 4.38E-06 | FALSE |
| GBA1 | 1-155235708-G-A | 1 | 155235708 | | G | A | Pathogenic | 1722541 | 2 | 1611508 | 1.24E-06 | FALSE |
| GBA1 | 1-155235757-C-T | 1 | 155235757 | rs1553217009 | C | T | Pathogenic | 496081 | 8 | 1613982 | 4.96E-06 | FALSE |
| GBA1 | 1-155235775-A-T | 1 | 155235775 | rs1557901552 | A | T | Pathogenic | 599275 | 1 | 1614214 | 6.19E-07 | FALSE |

| gnomAD (v4.1.0) pathogenic variant information for Parkinson's Disease genes | | | | | | | | | | | | |
|---|---|---|---|---|---|---|---|---|---|---|---|---|
| gene | gnomAD_ID | Chromosome | Position | rsIDs | Reference | Alternate | ClinVar_ClnSignificance | ClinVar_varID | allele_count | allele_number | allele_freq | in_gp2 |
| GBA1 | 1-155235780-G-A | 1 | 155235780 | rs76910485 | G | A | Pathogenic | 931820 | 3 | 1614220 | 1.86E-06 | FALSE |
| GBA1 | 1-155235810-C-T | 1 | 155235810 | rs1671711470 | C | T | Pathogenic | 918141 | 6 | 1614226 | 3.72E-06 | FALSE |
| GBA1 | 1-155236277-G-A | 1 | 155236277 | rs121908309 | G | A | Pathogenic | 4326 | 22 | 1614110 | 1.36E-05 | FALSE |
| GBA1 | 1-155236285-G-A | 1 | 155236285 | rs760307559 | G | A | Pathogenic | 918140 | 6 | 1614156 | 3.72E-06 | FALSE |
| GBA1 | 1-155236295-G-A | 1 | 155236295 | rs121908308 | G | A | Pathogenic | 813336 | 11 | 1614020 | 6.82E-06 | TRUE |
| GBA1 | 1-155236415-A-G | 1 | 155236415 | | A | G | Pathogenic | 2581173 | 1 | 1614166 | 6.20E-07 | FALSE |
| GBA1 | 1-155236439-CA-C | 1 | 155236439 | rs1553217314 | CA | C | Pathogenic | 496079 | 4 | 1614182 | 2.48E-06 | FALSE |
| GBA1 | 1-155237357-G-A | 1 | 155237357 | rs121908298 | G | A | Pathogenic | 4305 | 1 | 1613970 | 6.20E-07 | FALSE |
| GBA1 | 1-155237370-G-A | 1 | 155237370 | rs765633380 | G | A | Pathogenic | 813337 | 30 | 1613904 | 1.86E-05 | FALSE |
| GBA1 | 1-155237453-C-T | 1 | 155237453 | rs78973108 | C | T | Pathogenic | 4328 | 120 | 1613838 | 7.44E-05 | TRUE |
| GBA1 | 1-155237454-G-A | 1 | 155237454 | rs1553217626 | G | A | Pathogenic | 558787 | 1 | 1613960 | 6.20E-07 | FALSE |
| GBA1 | 1-155237520-C-T | 1 | 155237520 | | C | T | Pathogenic | 3064209 | 1 | 1614006 | 6.20E-07 | FALSE |
| GBA1 | 1-155238174-C-T | 1 | 155238174 | rs409652 | C | T | Pathogenic | 93459 | 50 | 1614020 | 3.10E-05 | FALSE |
| GBA1 | 1-155238194-C-T | 1 | 155238194 | rs74462743 | C | T | Pathogenic | 558788 | 13 | 1614154 | 8.05E-06 | FALSE |
| GBA1 | 1-155238206-A-C | 1 | 155238206 | rs381427 | A | C | Pathogenic | 928835 | 9 | 1613832 | 5.58E-06 | TRUE |
| GBA1 | 1-155238215-T-C | 1 | 155238215 | rs364897 | T | C | Pathogenic | 4314 | 100 | 1612400 | 6.20E-05 | TRUE |
| GBA1 | 1-155238242-C-T | 1 | 155238242 | rs867929413 | C | T | Pathogenic | 632834 | 4 | 1613234 | 2.48E-06 | FALSE |
| GBA1 | 1-155238260-G-C | 1 | 155238260 | rs1671872221 | G | C | Pathogenic | 996254 | 18 | 1609276 | 1.12E-05 | FALSE |
| GBA1 | 1-155238597-G-A | 1 | 155238597 | rs398123530 | G | A | Pathogenic | 93453 | 22 | 1613446 | 1.36E-05 | FALSE |
| GBA1 | 1-155238630-G-A | 1 | 155238630 | rs439898 | G | A | Pathogenic | 65570 | 43 | 1613512 | 2.66E-05 | TRUE |
| GBA1 | 1-155239736-G-A | 1 | 155239736 | rs1671974195 | G | A | Pathogenic | 984479 | 1 | 1614058 | 6.20E-07 | FALSE |
| GBA1 | 1-155239934-G-A | 1 | 155239934 | rs1141814 | G | A | Pathogenic | 4321 | 20 | 1614120 | 1.24E-05 | TRUE |
| GBA1 | 1-155239937-G-A | 1 | 155239937 | rs1671987417 | G | A | Pathogenic | 1321421 | 7 | 1614050 | 4.34E-06 | FALSE |
| GBA1 | 1-155239939-C-T | 1 | 155239939 | rs77829017 | C | T | Pathogenic | 4296 | 2 | 1614108 | 1.24E-06 | FALSE |
| GBA1 | 1-155239989-CG-C | 1 | 155239989 | rs1170895261 | CG | C | Pathogenic | 1321450 | 1 | 1613966 | 6.20E-07 | FALSE |
| GBA1 | 1-155240033-C-A | 1 | 155240033 | rs121908302 | C | A | Pathogenic | 4313 | 1 | 1614006 | 6.20E-07 | FALSE |
| GBA1 | 1-155240637-C-T | 1 | 155240637 | rs777383151 | C | T | Pathogenic | 974986 | 1 | 1613372 | 6.20E-07 | FALSE |
| GBA1 | 1-155240660-G-GC | 1 | 155240660 | rs387906315 | G | GC | Pathogenic | 4302 | 50 | 1613560 | 3.10E-05 | FALSE |
| PLA2G6 | 22-38112253-CTG-C | 22 | 38112253 | rs587784352 | CTG | C | Pathogenic | 159764 | 9 | 1613590 | 5.58E-06 | FALSE |
| PLA2G6 | 22-38112529-C-A | 22 | 38112529 | rs1296348337 | C | A | Pathogenic | 1012698 | 1 | 1555938 | 6.43E-07 | FALSE |
| PLA2G6 | 22-38113522-CA-C | 22 | 38113522 | rs1281282603 | CA | C | Pathogenic | 3214186 | 2 | 1613966 | 1.24E-06 | FALSE |
| PLA2G6 | 22-38113616-GACA-G | 22 | 38113616 | rs587784343 | GACA | G | Pathogenic | 6198 | 4 | 1613970 | 2.48E-06 | FALSE |
| PLA2G6 | 22-38115579-G-A | 22 | 38115579 | rs767689496 | G | A | Pathogenic | 2724426 | 3 | 1613568 | 1.86E-06 | FALSE |
| PLA2G6 | 22-38115587-G-C | 22 | 38115587 | | G | C | Pathogenic | 2170057 | 1 | 1613464 | 6.20E-07 | FALSE |
| PLA2G6 | 22-38115592-C-T | 22 | 38115592 | rs1318351016 | C | T | Pathogenic | 1686076 | 2 | 1613272 | 1.24E-06 | FALSE |
| PLA2G6 | 22-38115628-G-A | 22 | 38115628 | rs1484455290 | G | A | Pathogenic | 1028628 | 10 | 1607406 | 6.22E-06 | FALSE |
| PLA2G6 | 22-38115658-G-A | 22 | 38115658 | rs587784339 | G | A | Pathogenic | 159749 | 46 | 1544374 | 2.98E-05 | TRUE |
| PLA2G6 | 22-38115668-C-T | 22 | 38115668 | rs2145683127 | C | T | Pathogenic | 1180814 | 5 | 1600860 | 3.12E-06 | FALSE |
| PLA2G6 | 22-38115679-G-A | 22 | 38115679 | | G | A | Pathogenic | 2748479 | 1 | 1601154 | 6.25E-07 | FALSE |
| PLA2G6 | 22-38120835-C-A | 22 | 38120835 | rs777259654 | C | A | Pathogenic | 2862124 | 2 | 1613932 | 1.24E-06 | FALSE |
| PLA2G6 | 22-38123161-T-TG | 22 | 38123161 | rs1172241523 | T | TG | Pathogenic | 2737044 | 10 | 1550552 | 6.45E-06 | FALSE |
| PLA2G6 | 22-38123185-C-T | 22 | 38123185 | rs587784332 | C | T | Pathogenic | 379833 | 3 | 1552570 | 1.93E-06 | FALSE |
| PLA2G6 | 22-38126369-A-G | 22 | 38126369 | rs1352483031 | A | G | Pathogenic | 2412648 | 1 | 1611342 | 6.21E-07 | FALSE |
| PLA2G6 | 22-38126446-AG-A | 22 | 38126446 | rs587784329 | AG | A | Pathogenic | 159730 | 14 | 1612592 | 8.68E-06 | FALSE |
| PLA2G6 | 22-38128354-GA-G | 22 | 38128354 | rs1282370486 | GA | G | Pathogenic | 652932 | 6 | 1613734 | 3.72E-06 | FALSE |

**gnomAD (v4.1.0) pathogenic variant information for Parkinson's Disease genes**

| gene | gnomAD_ID | Chromosome | Position | rsIDs | Reference | Alternate | ClinVar_ClnSignificance | ClinVar_varID | allele_count | allele_number | allele_freq | in_gp2 |
|---|---|---|---|---|---|---|---|---|---|---|---|---|
| PLA2G6 | 22-38128402-CAG-C | 22 | 38128402 | rs1290554462 | CAG | C | Pathogenic | 2741274 | 4 | 1613996 | 2.48E-06 | FALSE |
| PLA2G6 | 22-38129453-C-A | 22 | 38129453 | rs761815070 | C | A | Pathogenic | 2581775 | 4 | 1598064 | 2.50E-06 | FALSE |
| PLA2G6 | 22-38129523-C-G | 22 | 38129523 | rs587784327 | C | G | Pathogenic | 2863736 | 2 | 1613964 | 1.24E-06 | FALSE |
| PLA2G6 | 22-38132869-C-T | 22 | 38132869 | rs1569263730 | C | T | Pathogenic | 561083 | 4 | 1552272 | 2.58E-06 | FALSE |
| PLA2G6 | 22-38132917-C-A | 22 | 38132917 | rs199935023 | C | A | Pathogenic | 30371 | 23 | 1557154 | 1.48E-05 | TRUE |
| PLA2G6 | 22-38135061-A-C | 22 | 38135061 | rs587784362 | A | C | Pathogenic | 159780 | 1 | 1602272 | 6.24E-07 | FALSE |
| PLA2G6 | 22-38140136-G-A | 22 | 38140136 | rs2088813126 | G | A | Pathogenic | 987078 | 1 | 1614122 | 6.20E-07 | FALSE |
| PLA2G6 | 22-38143177-CATCT-C | 22 | 38143177 | rs2089023572 | CATCT | C | Pathogenic | 986119 | 9 | 1614080 | 5.58E-06 | FALSE |
| PLA2G6 | 22-38169219-G-A | 22 | 38169219 | rs886039552 | G | A | Pathogenic | 265449 | 9 | 1611766 | 5.58E-06 | FALSE |
| PLA2G6 | 22-38169300-G-A | 22 | 38169300 | rs761956614 | G | A | Pathogenic | 2412654 | 1 | 1614184 | 6.20E-07 | FALSE |
| PLA2G6 | 22-38169318-G-A | 22 | 38169318 | rs200075782 | G | A | Pathogenic | 30370 | 58 | 1614026 | 3.59E-05 | TRUE |
| PLA2G6 | 22-38169426-T-C | 22 | 38169426 | rs1167198937 | T | C | Pathogenic | 2412655 | 2 | 1613872 | 1.24E-06 | FALSE |
| PINK1 | 1-20633561-C-T | 1 | 20633561 | rs1005937012 | C | T | Pathogenic | 661098 | 10 | 1182666 | 8.46E-06 | FALSE |
| PINK1 | 1-20633622-CCGGCCGGGCCTACC | 1 | 20633622 | rs1480758482 | CCGGCCGGGCC | C | Pathogenic | 1454290 | 3 | 1293628 | 2.32E-06 | FALSE |
| PINK1 | 1-20633820-GC-G | 1 | 20633820 | rs755000580 | GC | G | Pathogenic | 664114 | 2 | 1577720 | 1.27E-06 | FALSE |
| PINK1 | 1-20638052-GC-G | 1 | 20638052 | rs1557561340 | GC | G | Pathogenic | 581272 | 8 | 1614136 | 4.96E-06 | FALSE |
| PINK1 | 1-20638073-CG-C | 1 | 20638073 | rs756677845 | CG | C | Pathogenic | 189240 | 2 | 1613932 | 1.24E-06 | FALSE |
| PINK1 | 1-20639952-C-T | 1 | 20639952 | rs74315357 | C | T | Pathogenic | 2407 | 13 | 1612632 | 8.06E-06 | FALSE |
| PINK1 | 1-20639990-C-A | 1 | 20639990 | rs756783990 | C | A | Pathogenic | 631591 | 3 | 1602136 | 1.87E-06 | FALSE |
| PINK1 | 1-20645640-T-C | 1 | 20645640 | rs28940285 | T | C | Pathogenic | 2408 | 26 | 1614012 | 1.61E-05 | TRUE |
| PINK1 | 1-20648994-G-A | 1 | 20648994 | rs2053228483 | G | A | Pathogenic | 934238 | 1 | 1612924 | 6.20E-07 | FALSE |
| PINK1 | 1-20649054-G-A | 1 | 20649054 | rs74315356 | G | A | Pathogenic | 2406 | 8 | 1614040 | 4.96E-06 | FALSE |
| PINK1 | 1-20649070-GC-G | 1 | 20649070 | rs775479526 | GC | G | Pathogenic | 1399771 | 30 | 1614094 | 1.86E-05 | FALSE |
| PINK1 | 1-20649109-C-T | 1 | 20649109 | rs45539432 | C | T | Pathogenic | 2415 | 66 | 1614232 | 4.09E-05 | TRUE |
| PINK1 | 1-20649217-C-T | 1 | 20649217 | rs34208370 | C | T | Pathogenic | 431963 | 101 | 1614054 | 6.26E-05 | TRUE |
| PINK1 | 1-20649232-G-A | 1 | 20649232 | rs2053233432 | G | A | Pathogenic | 961640 | 4 | 1613880 | 2.48E-06 | FALSE |
| PINK1 | 1-20650541-C-CCAA | 1 | 20650541 | rs750664040 | C | CCAA | Pathogenic | 2410 | 8 | 1614092 | 4.96E-06 | FALSE |
| PARK7 | 1-7962867-C-T | 1 | 7962867 | rs374429170 | C | T | Pathogenic | 1399835 | 9 | 1612268 | 5.58E-06 | FALSE |
| PARK7 | 1-7965336-G-GT | 1 | 7965336 | rs781600849 | G | GT | Pathogenic | 573287 | 32 | 1613906 | 1.98E-05 | FALSE |
| PARK7 | 1-7965425-G-C | 1 | 7965425 | rs74315353 | G | C | Pathogenic | 7067 | 3 | 1613528 | 1.86E-06 | FALSE |
| PARK7 | 1-7970963-G-A | 1 | 7970963 | rs1252815484 | G | A | Pathogenic | 1334459 | 2 | 1614192 | 1.24E-06 | FALSE |
| LRRK2 | 12-40299125-A-G | 12 | 40299125 | rs34805604 | A | G | Pathogenic | 1939 | 2 | 1612972 | 1.24E-06 | FALSE |
| LRRK2 | 12-40310434-C-G | 12 | 40310434 | rs33939927 | C | G | Pathogenic | 1936 | 5 | 1612956 | 3.10E-06 | TRUE |
| LRRK2 | 12-40310434-C-T | 12 | 40310434 | rs33939927 | C | T | Pathogenic | 1938 | 39 | 1612956 | 2.42E-05 | TRUE |
| LRRK2 | 12-40320129-C-T | 12 | 40320129 | | C | T | Pathogenic | 2572065 | 2 | 1611666 | 1.24E-06 | FALSE |
| PRKN | 6-161350163-C-T | 6 | 161350163 | rs961239925 | C | T | Pathogenic | 1459191 | 4 | 1613678 | 2.48E-06 | FALSE |
| PRKN | 6-161350214-G-C | 6 | 161350214 | rs765860776 | G | C | Pathogenic | 2136491 | 5 | 1608138 | 3.11E-06 | TRUE |
| PRKN | 6-161548965-GA-G | 6 | 161548965 | rs1562519380 | GA | G | Pathogenic | 1456905 | 8 | 1614124 | 4.96E-06 | FALSE |
| PRKN | 6-161569417-C-G | 6 | 161569417 | rs772074730 | C | G | Pathogenic | 1068478 | 2 | 1613248 | 1.24E-06 | FALSE |
| PRKN | 6-161785793-C-G | 6 | 161785793 | rs751037529 | C | G | Pathogenic | 409266 | 7 | 1614020 | 4.34E-06 | TRUE |
| PRKN | 6-161785839-A-T | 6 | 161785839 | rs377554392 | A | T | Pathogenic | 805244 | 13 | 1614148 | 8.05E-06 | TRUE |
| PRKN | 6-161973317-G-C | 6 | 161973317 | rs137853054 | G | C | Pathogenic | 7036 | 1 | 1610416 | 6.21E-07 | FALSE |
| PRKN | 6-161973401-C-T | 6 | 161973401 | rs137853058 | C | T | Pathogenic | 7046 | 7 | 1610136 | 4.35E-06 | FALSE |
| PRKN | 6-162054107-C-CT | 6 | 162054107 | rs1231455463 | C | CT | Pathogenic | 2174566 | 1 | 1609692 | 6.21E-07 | FALSE |
| PRKN | 6-162443314-A-T | 6 | 162443314 | rs137853059 | A | T | Pathogenic | 7047 | 33 | 1612238 | 2.05E-05 | TRUE |

**gnomAD (v4.1.0) pathogenic variant information for Parkinson's Disease genes**

| gene | gnomAD_ID | Chromosome | Position | rsIDs | Reference | Alternate | ClinVar_ClnSignificance | ClinVar_varID | allele_count | allele_number | allele_freq | in_gp2 |
|---|---|---|---|---|---|---|---|---|---|---|---|---|
| PRKN | 6-162443325-AT-A | 6 | 162443325 | rs754809877 | AT | A | Pathogenic | 536457 | 449 | 1612394 | 0.000278467917 | FALSE |
| PRKN | 6-162443356-C-G | 6 | 162443356 | rs368134308 | C | G | Pathogenic | 644125 | 67 | 1613226 | 4.15E-05 | TRUE |
| PRKN | 6-162443378-CCT-C | 6 | 162443378 | rs55777503 | CCT | C | Pathogenic | 425403 | 543 | 1613910 | 0.000336449987 | FALSE |
| PRKN | 6-162443384-G-A | 6 | 162443384 | rs770591350 | G | A | Pathogenic | 853293 | 10 | 1613756 | 6.20E-06 | FALSE |
| POLG | 15-89317374-A-G | 15 | 89317374 | rs1335880349 | A | G | Pathogenic | 597808 | 6 | 1613830 | 3.72E-06 | FALSE |
| POLG | 15-89317375-C-T | 15 | 89317375 | rs1326779034 | C | T | Pathogenic | 2677947 | 1 | 1613940 | 6.20E-07 | FALSE |
| POLG | 15-89317389-G-GTATC | 15 | 89317389 | rs2055307723 | G | GTATC | Pathogenic | 2677952 | 8 | 1613984 | 4.96E-06 | FALSE |
| POLG | 15-89317448-TC-T | 15 | 89317448 | | TC | T | Pathogenic | 2024844 | 1 | 1614094 | 6.20E-07 | FALSE |
| POLG | 15-89317469-C-T | 15 | 89317469 | rs1131691575 | C | T | Pathogenic | 426100 | 4 | 1613976 | 2.48E-06 | FALSE |
| POLG | 15-89318540-C-A | 15 | 89318540 | | C | A | Pathogenic | 3013692 | 1 | 1612918 | 6.20E-07 | FALSE |
| POLG | 15-89318694-TGTAA-T | 15 | 89318694 | rs1442498340 | TGTAA | T | Pathogenic | 1457717 | 1 | 1614068 | 6.20E-07 | FALSE |
| POLG | 15-89318947-GAGGGCTCC-G | 15 | 89318947 | | GAGGGCTCC | G | Pathogenic | 2645687 | 1 | 1614088 | 6.20E-07 | FALSE |
| POLG | 15-89318963-G-A | 15 | 89318963 | rs767708989 | G | A | Pathogenic | 619310 | 5 | 1613954 | 3.10E-06 | FALSE |
| POLG | 15-89318986-G-A | 15 | 89318986 | rs267606959 | G | A | Pathogenic | 13516 | 18 | 1614078 | 1.12E-05 | FALSE |
| POLG | 15-89319044-CTG-C | 15 | 89319044 | rs1332921412 | CTG | C | Pathogenic | 817632 | 2 | 1614062 | 1.24E-06 | FALSE |
| POLG | 15-89319048-G-GC | 15 | 89319048 | rs1447799185 | G | GC | Pathogenic | 458711 | 2 | 1614028 | 1.24E-06 | FALSE |
| POLG | 15-89319225-T-A | 15 | 89319225 | rs778573169 | T | A | Pathogenic | 587863 | 31 | 1614058 | 1.92E-05 | TRUE |
| POLG | 15-89319229-TC-T | 15 | 89319229 | rs2055358006 | TC | T | Pathogenic | 872916 | 1 | 1614144 | 6.20E-07 | FALSE |
| POLG | 15-89319265-G-A | 15 | 89319265 | rs1567185770 | G | A | Pathogenic | 619307 | 1 | 1614180 | 6.20E-07 | FALSE |
| POLG | 15-89319274-TCCAGCCACCCTCA | 15 | 89319274 | rs886041276 | TCCAGCCACCC | T | Pathogenic | 279948 | 1 | 1614148 | 6.20E-07 | FALSE |
| POLG | 15-89319318-ACCAGCCACTCG-A | 15 | 89319318 | rs2055359441 | ACCAGCCACTC | A | Pathogenic | 1449732 | 1 | 1599762 | 6.25E-07 | FALSE |
| POLG | 15-89319349-AC-A | 15 | 89319349 | rs1314787391 | AC | A | Pathogenic | 2866014 | 1 | 1613918 | 6.20E-07 | FALSE |
| POLG | 15-89320883-T-C | 15 | 89320883 | rs113994099 | T | C | Pathogenic | 13495 | 1 | 1613974 | 6.20E-07 | FALSE |
| POLG | 15-89320919-C-T | 15 | 89320919 | rs1567186613 | C | T | Pathogenic | 619302 | 1 | 1613964 | 6.20E-07 | FALSE |
| POLG | 15-89320953-G-A | 15 | 89320953 | rs121918048 | G | A | Pathogenic | 13500 | 3 | 1614004 | 1.86E-06 | FALSE |
| POLG | 15-89321184-T-TC | 15 | 89321184 | rs1283198587 | T | TC | Pathogenic | 619306 | 4 | 1614062 | 2.48E-06 | FALSE |
| POLG | 15-89321242-C-A | 15 | 89321242 | rs121918047 | C | A | Pathogenic | 13504 | 3 | 1614188 | 1.86E-06 | FALSE |
| POLG | 15-89321743-T-C | 15 | 89321743 | rs121918050 | T | C | Pathogenic | 13506 | 3 | 1611710 | 1.86E-06 | FALSE |
| POLG | 15-89321780-G-A | 15 | 89321780 | rs144500145 | G | A | Pathogenic | 206528 | 179 | 1613984 | 0.000110905684 | TRUE |
| POLG | 15-89321783-T-C | 15 | 89321783 | rs775445970 | T | C | Pathogenic | 619395 | 5 | 1613898 | 3.10E-06 | FALSE |
| POLG | 15-89321792-C-T | 15 | 89321792 | rs113994098 | C | T | Pathogenic | 13502 | 498 | 1614136 | 0.000308524188 | TRUE |
| POLG | 15-89321961-C-T | 15 | 89321961 | rs1567187326 | C | T | Pathogenic | 619400 | 5 | 1614106 | 3.10E-06 | FALSE |
| POLG | 15-89321961-C-A | 15 | 89321961 | | C | A | Pathogenic | 2860133 | 1 | 1614106 | 6.20E-07 | FALSE |
| POLG | 15-89322748-C-G | 15 | 89322748 | rs796052887 | C | G | Pathogenic | 2736252 | 3 | 1613940 | 1.86E-06 | FALSE |
| POLG | 15-89322856-AG-A | 15 | 89322856 | | AG | A | Pathogenic | 2760375 | 1 | 1613918 | 6.20E-07 | FALSE |
| POLG | 15-89323452-G-T | 15 | 89323452 | rs750514687 | G | T | Pathogenic | 1390135 | 1 | 1614106 | 6.20E-07 | FALSE |
| POLG | 15-89323829-G-A | 15 | 89323829 | rs1254855971 | G | A | Pathogenic | 619398 | 4 | 1613492 | 2.48E-06 | FALSE |
| POLG | 15-89323847-G-A | 15 | 89323847 | rs867038717 | G | A | Pathogenic | 381519 | 4 | 1613918 | 2.48E-06 | FALSE |
| POLG | 15-89324138-AG-A | 15 | 89324138 | rs1567188632 | AG | A | Pathogenic | 619396 | 10 | 1614102 | 6.20E-06 | FALSE |
| POLG | 15-89325452-G-T | 15 | 89325452 | rs1465650547 | G | T | Pathogenic | 969298 | 2 | 1598360 | 1.25E-06 | FALSE |
| POLG | 15-89325520-G-A | 15 | 89325520 | rs121918046 | G | A | Pathogenic | 13499 | 7 | 1611770 | 4.34E-06 | FALSE |
| POLG | 15-89325539-C-T | 15 | 89325539 | | C | T | Pathogenic | 2748199 | 1 | 1613248 | 6.20E-07 | FALSE |
| POLG | 15-89325610-G-A | 15 | 89325610 | rs139717885 | G | A | Pathogenic | 381520 | 23 | 1613526 | 1.43E-05 | TRUE |
| POLG | 15-89325616-G-A | 15 | 89325616 | rs2152065937 | G | A | Pathogenic | 1421810 | 1 | 1613514 | 6.20E-07 | FALSE |
| POLG | 15-89325679-G-A | 15 | 89325679 | rs774474723 | G | A | Pathogenic | 1452045 | 13 | 1605794 | 8.10E-06 | TRUE |

**gnomAD (v4.1.0) pathogenic variant information for Parkinson's Disease genes**

| gene | gnomAD_ID | Chromosome | Position | rsIDs | Reference | Alternate | ClinVar_ClnSignificance | ClinVar_varID | allele_count | allele_number | allele_freq | in_gp2 |
|---|---|---|---|---|---|---|---|---|---|---|---|---|
| POLG | 15-89326934-AG-A | 15 | 89326934 | rs1567191094 | AG | A | Pathogenic | 619448 | 1 | 1614088 | 6.20E-07 | FALSE |
| POLG | 15-89327040-C-T | 15 | 89327040 | rs2055531147 | C | T | Pathogenic | 1455434 | 1 | 1614216 | 6.19E-07 | FALSE |
| POLG | 15-89327046-C-T | 15 | 89327046 | | C | T | Pathogenic | 2810586 | 2 | 1614234 | 1.24E-06 | FALSE |
| POLG | 15-89327062-AC-A | 15 | 89327062 | rs2055531567 | AC | A | Pathogenic | 2843895 | 1 | 1614072 | 6.20E-07 | FALSE |
| POLG | 15-89327166-C-T | 15 | 89327166 | rs771623994 | C | T | Pathogenic | 280017 | 14 | 1614140 | 8.67E-06 | TRUE |
| POLG | 15-89327201-C-T | 15 | 89327201 | rs113994095 | C | T | Pathogenic | 13496 | 1637 | 1614256 | 0.001014089463 | TRUE |
| POLG | 15-89327244-A-C | 15 | 89327244 | | A | C | Pathogenic | 2677961 | 2 | 1614190 | 1.24E-06 | FALSE |
| POLG | 15-89327306-CA-C | 15 | 89327306 | | CA | C | Pathogenic | 2865640 | 1 | 1614050 | 6.20E-07 | FALSE |
| POLG | 15-89327311-A-G | 15 | 89327311 | rs1567191474 | A | G | Pathogenic | 619407 | 2 | 1613990 | 1.24E-06 | FALSE |
| POLG | 15-89327328-CAG-C | 15 | 89327328 | rs796052908 | CAG | C | Pathogenic | 206608 | 5 | 1613734 | 3.10E-06 | FALSE |
| POLG | 15-89327351-T-C | 15 | 89327351 | rs2055536585 | T | C | Pathogenic | 933883 | 1 | 1612794 | 6.20E-07 | FALSE |
| POLG | 15-89328735-G-A | 15 | 89328735 | rs960142425 | G | A | Pathogenic | 619394 | 4 | 1613996 | 2.48E-06 | FALSE |
| POLG | 15-89328781-CT-C | 15 | 89328781 | rs763418954 | CT | C | Pathogenic | 596511 | 12 | 1614042 | 7.43E-06 | FALSE |
| POLG | 15-89329041-G-A | 15 | 89329041 | rs886041592 | G | A | Pathogenic | 280375 | 7 | 1613060 | 4.34E-06 | TRUE |
| POLG | 15-89329059-C-T | 15 | 89329059 | rs749799663 | C | T | Pathogenic | 1425375 | 7 | 1612940 | 4.34E-06 | FALSE |
| POLG | 15-89330113-G-A | 15 | 89330113 | rs1057517803 | G | A | Pathogenic | 372472 | 5 | 1613966 | 3.10E-06 | FALSE |
| POLG | 15-89330231-C-T | 15 | 89330231 | rs1567192879 | C | T | Pathogenic | 619303 | 2 | 1613190 | 1.24E-06 | FALSE |
| POLG | 15-89330241-C-T | 15 | 89330241 | rs113994093 | C | T | Pathogenic | 21319 | 14 | 1612638 | 8.68E-06 | FALSE |
| POLG | 15-89333526-G-A | 15 | 89333526 | | G | A | Pathogenic | 2000637 | 1 | 1612882 | 6.20E-07 | FALSE |
| POLG | 15-89333553-G-A | 15 | 89333553 | rs202039305 | G | A | Pathogenic | 279988 | 3 | 1612020 | 1.86E-06 | FALSE |
| POLG | 15-89333619-G-A | 15 | 89333619 | rs2055628382 | G | A | Pathogenic | 2707771 | 1 | 1601374 | 6.24E-07 | FALSE |
| POLG | 15-89333680-C-T | 15 | 89333680 | rs1021719232 | C | T | Pathogenic | 694427 | 2 | 1548958 | 1.29E-06 | FALSE |
| POLG | 15-89333747-C-G | 15 | 89333747 | rs121918045 | C | G | Pathogenic | 13498 | 1 | 1535292 | 6.51E-07 | FALSE |
| DNAJC6 | 1-65366107-C-T | 1 | 65366107 | rs864622011 | C | T | Pathogenic | 219301 | 3 | 1613868 | 1.86E-06 | FALSE |
| DNAJC6 | 1-65385899-C-T | 1 | 65385899 | rs1645867120 | C | T | Pathogenic | 976692 | 4 | 1600054 | 2.50E-06 | FALSE |
| SYNJ1 | 21-32638958-G-A | 21 | 32638958 | rs747261340 | G | A | Pathogenic | 1076901 | 11 | 1613954 | 6.82E-06 | FALSE |
| SYNJ1 | 21-32643449-C-CT | 21 | 32643449 | rs2145756643 | C | CT | Pathogenic | 1070684 | 2 | 1613586 | 1.24E-06 | FALSE |
| SYNJ1 | 21-32645791-T-C | 21 | 32645791 | rs1057524880 | T | C | Pathogenic | 393360 | 1 | 1482182 | 6.75E-07 | FALSE |
| SYNJ1 | 21-32646432-G-A | 21 | 32646432 | rs1373545506 | G | A | Pathogenic | 978699 | 1 | 1614094 | 6.20E-07 | FALSE |
| SYNJ1 | 21-32646513-TG-T | 21 | 32646513 | rs1230133310 | TG | T | Pathogenic | 848199 | 1 | 1613948 | 6.20E-07 | FALSE |
| SYNJ1 | 21-32656686-CCTTAT-C | 21 | 32656686 | rs778394516 | CCTTAT | C | Pathogenic | 570902 | 5 | 1610262 | 3.11E-06 | FALSE |
| SYNJ1 | 21-32665963-G-A | 21 | 32665963 | | G | A | Pathogenic | 1954917 | 5 | 1609376 | 3.11E-06 | FALSE |
| SYNJ1 | 21-32678675-G-A | 21 | 32678675 | | G | A | Pathogenic | 2936446 | 1 | 1609890 | 6.21E-07 | FALSE |
| SYNJ1 | 21-32685772-T-TA | 21 | 32685772 | rs1419316294 | T | TA | Pathogenic | 1074545 | 10 | 1606784 | 6.22E-06 | FALSE |
| SYNJ1 | 21-32694269-G-A | 21 | 32694269 | rs1160469053 | G | A | Pathogenic | 1456091 | 4 | 1567130 | 2.55E-06 | FALSE |
| SYNJ1 | 21-32695106-C-T | 21 | 32695106 | rs398122403 | C | T | Pathogenic | 88844 | 17 | 1614092 | 1.05E-05 | FALSE |
| SYNJ1 | 21-32695131-G-A | 21 | 32695131 | rs756965178 | G | A | Pathogenic | 1323673 | 7 | 1613898 | 4.34E-06 | FALSE |
| SYNJ1 | 21-32700028-G-A | 21 | 32700028 | rs2042344677 | G | A | Pathogenic | 1206765 | 4 | 1613928 | 2.48E-06 | FALSE |
| SYNJ1 | 21-32726907-C-A | 21 | 32726907 | rs1040540690 | C | A | Pathogenic | 1457606 | 7 | 1613892 | 4.34E-06 | TRUE |
| SYNJ1 | 21-32728027-C-CAT | 21 | 32728027 | rs1227986180 | C | CAT | Pathogenic | 1069960 | 1 | 1535332 | 6.51E-07 | FALSE |
| SYNJ1 | 21-32728032-CT-C | 21 | 32728032 | | CT | C | Pathogenic | 2945224 | 1 | 1533782 | 6.52E-07 | FALSE |
| VPS13C | 15-61876978-C-A | 15 | 61876978 | rs751054856 | C | A | Pathogenic | 3006540 | 10 | 1599020 | 6.25E-06 | FALSE |
| VPS13C | 15-61878689-C-A | 15 | 61878689 | rs199723460 | C | A | Pathogenic | 1323753 | 1 | 1611370 | 6.21E-07 | FALSE |
| VPS13C | 15-61881570-G-A | 15 | 61881570 | rs1229922592 | G | A | Pathogenic | 2993124 | 5 | 1606698 | 3.11E-06 | FALSE |
| VPS13C | 15-61884244-GT-G | 15 | 61884244 | rs775841187 | GT | G | Pathogenic | 2662741 | 52 | 1610372 | 3.23E-05 | FALSE |

**gnomAD (v4.1.0) pathogenic variant information for Parkinson's Disease genes**

| gene | gnomAD_ID | Chromosome | Position | rsIDs | Reference | Alternate | ClinVar_ClnSignificance | ClinVar_varID | allele_count | allele_number | allele_freq | in_gp2 |
|---|---|---|---|---|---|---|---|---|---|---|---|---|
| VPS13C | 15-61890331-G-A | 15 | 61890331 | rs1387456031 | G | A | Pathogenic | 2984894 | 7 | 1613896 | 4.34E-06 | FALSE |
| VPS13C | 15-61920161-A-AT | 15 | 61920161 | rs1315150327 | A | AT | Pathogenic | 1965239 | 7 | 1613480 | 4.34E-06 | FALSE |
| VPS13C | 15-61951850-A-AT | 15 | 61951850 | | A | AT | Pathogenic | 2822600 | 4 | 1611338 | 2.48E-06 | FALSE |
| VPS13C | 15-61961648-G-T | 15 | 61961648 | rs1180158172 | G | T | Pathogenic | 3238850 | 3 | 1613706 | 1.86E-06 | FALSE |
| VPS13C | 15-62023829-CT-C | 15 | 62023829 | rs1456557102 | CT | C | Pathogenic | 2724228 | 10 | 1610480 | 6.21E-06 | FALSE |
| VPS13C | 15-62033477-G-A | 15 | 62033477 | rs778239562 | G | A | Pathogenic | 1941006 | 34 | 1604990 | 2.12E-05 | FALSE |
| VPS13C | 15-62035047-AT-A | 15 | 62035047 | rs761323769 | AT | A | Pathogenic | 3000824 | 9 | 1602338 | 5.62E-06 | FALSE |
| HTRA2 | 2-74530250-C-T | 2 | 74530250 | rs1675486182 | C | T | Pathogenic | 1424738 | 7 | 1609358 | 4.35E-06 | FALSE |
| HTRA2 | 2-74532641-C-T | 2 | 74532641 | rs1407675367 | C | T | Pathogenic | 1455851 | 13 | 1613560 | 8.06E-06 | FALSE |
| HTRA2 | 2-74532714-G-A | 2 | 74532714 | rs767006508 | G | A | Pathogenic | 372209 | 14 | 1613412 | 8.68E-06 | FALSE |
| TMEM230 | 20-5100921-C-A | 20 | 5100921 | rs764786986 | C | A | Pathogenic | 243014 | 5 | 1613958 | 3.10E-06 | TRUE |
| UCHL1 | 4-41256996-A-C | 4 | 41256996 | rs397515634 | A | C | Pathogenic | 88635 | 2 | 1614182 | 1.24E-06 | FALSE |
| UCHL1 | 4-41257706-C-CGCT | 4 | 41257706 | rs749368841 | C | CGCT | Pathogenic | 2077519 | 3 | 1580822 | 1.90E-06 | FALSE |
| UCHL1 | 4-41261751-CAG-C | 4 | 41261751 | rs1310363710 | CAG | C | Pathogenic | 3068443 | 1 | 1613326 | 6.20E-07 | FALSE |
| UCHL1 | 4-41264108-C-T | 4 | 41264108 | rs2154087267 | C | T | Pathogenic | 1380167 | 1 | 1614208 | 6.19E-07 | FALSE |
