## Supplementary Table 5 for "Parkinson’s Disease Pathogenic Variants: Cross-Ancestry Analysis and Microarray Data Validation"

**All pathogenic variants per ancestry with freq (overall and case control)**

| | | | AAC | | | AFR | | | AJ | | | AMR | | | CAH | | | CAS | | | EAS | | | EUR | | | FIN | | | MDE | | | SAS | | |
|---|---|---|---|---|---|---|---|---|---|---|---|---|---|---|---|---|---|---|---|---|---|---|---|---|---|---|---|---|---|---|---|---|---|---|---|
| GENE | rsID | snpID | MAF | MAF_A | MAF_U | MAF | MAF_A | MAF_U | MAF | MAF_A | MAF_U | MAF | MAF_A | MAF_U | MAF | MAF_A | MAF_U | MAF | MAF_A | MAF_U | MAF | MAF_A | MAF_U | MAF | MAF_A | MAF_U | MAF | MAF_A | MAF_U | MAF | MAF_A | MAF_U | MAF | MAF_A | MAF_U |
| LRRK2 | rs33939927 | chr12:40310434:C:T | NA | NA | NA | NA | NA | NA | NA | NA | NA | NA | NA | NA | NA | NA | NA | 0.0005537 | 0.0009058 | <1e-6 | NA | NA | NA | 0.0002189 | 0.0003303 | 5.43E-05 | NA | NA | NA | 0.003448 | 0.006452 | <1e-6 | NA | NA | NA |
| POLG | rs778573169 | chr15:89319225:T:A | NA | NA | NA | NA | NA | NA | NA | NA | NA | NA | NA | NA | NA | NA | NA | NA | NA | NA | NA | NA | NA | 3.86E-05 | 4.72E-05 | <1e-6 | NA | NA | NA | NA | NA | NA | NA | NA | NA |
| POLG | rs144500145 | chr15:89321780:G:A | NA | NA | NA | NA | NA | NA | NA | NA | NA | NA | NA | NA | NA | NA | NA | NA | NA | NA | NA | NA | NA | 9.01E-05 | 9.44E-05 | 0.0001628 | NA | NA | NA | NA | NA | NA | NA | NA | NA |
| POLG | rs113994098 | chr15:89321792:C:T | <1e-6 | <1e-6 | <1e-6 | 0.0001893 | <1e-6 | 0.000298 | NA | NA | NA | NA | NA | NA | NA | NA | NA | NA | NA | NA | NA | NA | NA | 0.0002833 | 0.0002831 | 0.0002171 | NA | NA | NA | NA | NA | NA | NA | NA | NA |
| POLG | rs139717885 | chr15:89325610:G:A | NA | NA | NA | <1e-6 | <1e-6 | <1e-6 | NA | NA | NA | NA | NA | NA | NA | NA | NA | NA | NA | NA | NA | NA | NA | NA | NA | NA | NA | NA | NA | NA | NA | NA | NA | NA | NA |
| POLG | rs774474723 | chr15:89325679:G:A | NA | NA | NA | NA | NA | NA | NA | NA | NA | NA | NA | NA | NA | NA | NA | NA | NA | NA | NA | NA | NA | 2.58E-05 | <1e-6 | 5.43E-05 | NA | NA | NA | NA | NA | NA | NA | NA | NA |
| POLG | rs771623994 | chr15:89327166:C:T | NA | NA | NA | 0.0001897 | 0.0005313 | <1e-6 | NA | NA | NA | NA | NA | NA | NA | NA | NA | NA | NA | NA | NA | NA | NA | NA | NA | NA | NA | NA | NA | NA | NA | NA | NA | NA | NA |
| POLG | rs113994095 | chr15:89327201:C:T | 0.00045 | <1e-6 | 0.0006242 | 0.0001892 | <1e-6 | 0.0002978 | <1e-6 | <1e-6 | <1e-6 | NA | NA | NA | NA | NA | NA | NA | NA | NA | NA | NA | NA | 0.0007355 | 0.000804 | 0.0005438 | NA | NA | NA | NA | NA | NA | NA | NA | NA |
| POLG | rs886041592 | chr15:89329041:G:A | NA | NA | NA | NA | NA | NA | NA | NA | NA | NA | NA | NA | NA | NA | NA | NA | NA | NA | NA | NA | NA | 2.58E-05 | 4.72E-05 | <1e-6 | NA | NA | NA | NA | NA | NA | NA | NA | NA |
| GBA | rs80356771 | chr1:155235196:G:A | NA | NA | NA | NA | NA | NA | NA | NA | NA | NA | NA | NA | NA | NA | NA | NA | NA | NA | NA | NA | NA | 0.0003479 | 0.0005904 | 0.0001086 | NA | NA | NA | NA | NA | NA | NA | NA | NA |
| GBA | rs121908308 | chr1:155236295:G:A | NA | NA | NA | NA | NA | NA | NA | NA | NA | NA | NA | NA | 0.0005875 | 0.0009524 | <1e-6 | NA | NA | NA | NA | NA | NA | NA | NA | NA | NA | NA | NA | NA | NA | NA | NA | NA | NA |
| GBA | rs78973108 | chr1:155237453:C:T | NA | NA | NA | NA | NA | NA | NA | NA | NA | NA | NA | NA | NA | NA | NA | NA | NA | NA | NA | NA | NA | 0.000206 | 0.0002832 | 5.43E-05 | NA | NA | NA | NA | NA | NA | NA | NA | NA |
| GBA | rs381427 | chr1:155238206:A:C | NA | NA | NA | <1e-6 | <1e-6 | <1e-6 | NA | NA | NA | NA | NA | NA | NA | NA | NA | NA | NA | NA | NA | NA | NA | NA | NA | NA | NA | NA | NA | NA | NA | NA | NA | NA | NA |
| GBA | rs364897 | chr1:155238215:T:C | NA | NA | NA | 0.0001892 | 0.0005308 | <1e-6 | NA | NA | NA | NA | NA | NA | NA | NA | NA | NA | NA | NA | NA | NA | NA | <1e-6 | <1e-6 | <1e-6 | NA | NA | NA | NA | NA | NA | NA | NA | NA |
| GBA | rs439898 | chr1:155238630:G:A | NA | NA | NA | NA | NA | NA | NA | NA | NA | NA | NA | NA | NA | NA | NA | NA | NA | NA | NA | NA | NA | 6.44E-05 | 9.44E-05 | <1e-6 | NA | NA | NA | NA | NA | NA | NA | NA | NA |
| GBA | rs1141814 | chr1:155239934:G:A | NA | NA | NA | NA | NA | NA | NA | NA | NA | NA | NA | NA | NA | NA | NA | NA | NA | NA | NA | NA | NA | NA | NA | NA | NA | NA | NA | 0.001721 | 0.003215 | <1e-6 | NA | NA | NA |
| ATP13A2 | rs144701072 | chr1:16988455:C:T | NA | NA | NA | 0.0001892 | 0.0005308 | <1e-6 | NA | NA | NA | NA | NA | NA | NA | NA | NA | NA | NA | NA | NA | NA | NA | NA | NA | NA | NA | NA | NA | NA | NA | NA | NA | NA | NA |
| ATP13A2 | rs866035312 | chr1:16989961:G:A | NA | NA | NA | NA | NA | NA | NA | NA | NA | <1e-6 | <1e-6 | <1e-6 | NA | NA | NA | NA | NA | NA | NA | NA | NA | NA | NA | NA | NA | NA | NA | NA | NA | NA | NA | NA | NA |
| ATP13A2 | rs773246271 | chr1:16992345:G:A | NA | NA | NA | NA | NA | NA | NA | NA | NA | NA | NA | NA | NA | NA | NA | NA | NA | NA | NA | NA | NA | 2.58E-05 | 4.72E-05 | <1e-6 | NA | NA | NA | NA | NA | NA | NA | NA | NA |
| ATP13A2 | rs1303653650 | chr1:16996059:G:A | NA | NA | NA | NA | NA | NA | NA | NA | NA | NA | NA | NA | NA | NA | NA | NA | NA | NA | 0.0001935 | 0.0001878 | 0.0002032 | <1e-6 | <1e-6 | <1e-6 | NA | NA | NA | NA | NA | NA | NA | NA | NA |
| ATP13A2 | rs373607247 | chr1:17005450:C:T | NA | NA | NA | NA | NA | NA | NA | NA | NA | NA | NA | NA | NA | NA | NA | NA | NA | NA | NA | NA | NA | <1e-6 | <1e-6 | <1e-6 | NA | NA | NA | NA | NA | NA | NA | NA | NA |
| PINK1 | rs28940285 | chr1:20645640:T:C | NA | NA | NA | NA | NA | NA | NA | NA | NA | NA | NA | NA | NA | NA | NA | NA | NA | NA | 0.001941 | 0.003394 | 0.0004072 | NA | NA | NA | NA | NA | NA | NA | NA | NA | NA | NA | NA |
| PINK1 | rs45539432 | chr1:20649109:C:T | NA | NA | NA | NA | NA | NA | NA | NA | NA | NA | NA | NA | NA | NA | NA | NA | NA | NA | NA | NA | NA | 0.0002704 | 0.0003775 | 0.0001628 | NA | NA | NA | 0.0008606 | 0.001608 | <1e-6 | NA | NA | NA |
| PINK1 | rs34208370 | chr1:20649217:C:T | NA | NA | NA | NA | NA | NA | NA | NA | NA | NA | NA | NA | NA | NA | NA | NA | NA | NA | 0.0005812 | 0.0007522 | 0.0004067 | 3.86E-05 | 2.36E-05 | 0.0001086 | <1e-6 | <1e-6 | <1e-6 | NA | NA | NA | NA | NA | NA |
| TMEM230 | rs764786986 | chr20:5100921:C:A | NA | NA | NA | NA | NA | NA | NA | NA | NA | NA | NA | NA | NA | NA | NA | NA | NA | NA | NA | NA | NA | <1e-6 | <1e-6 | <1e-6 | NA | NA | NA | NA | NA | NA | NA | NA | NA |
| SYNJ1 | rs1040540690 | chr21:32726907:C:A | NA | NA | NA | 0.0001899 | 0.0005336 | <1e-6 | NA | NA | NA | NA | NA | NA | NA | NA | NA | NA | NA | NA | NA | NA | NA | NA | NA | NA | NA | NA | NA | NA | NA | NA | NA | NA | NA |
| PLA2G6 | rs587784339 | chr22:38115658:G:A | NA | NA | NA | NA | NA | NA | NA | NA | NA | 0.000774 | <1e-6 | 0.003226 | NA | NA | NA | NA | NA | NA | NA | NA | NA | 2.58E-05 | 4.72E-05 | <1e-6 | NA | NA | NA | NA | NA | NA | NA | NA | NA |
| PLA2G6 | rs199935023 | chr22:38132917:C:A | NA | NA | NA | NA | NA | NA | NA | NA | NA | NA | NA | NA | NA | NA | NA | NA | NA | NA | 0.0001937 | 0.0001881 | 0.0002033 | NA | NA | NA | NA | NA | NA | NA | NA | NA | NA | NA | NA |
| PLA2G6 | rs200075782 | chr22:38169318:G:A | NA | NA | NA | NA | NA | NA | NA | NA | NA | NA | NA | NA | NA | NA | NA | 0.0005537 | 0.0009058 | <1e-6 | NA | NA | NA | NA | NA | NA | NA | NA | NA | NA | NA | NA | NA | NA | NA |
| PRKN | rs765860776 | chr6:161350214:G:C | NA | NA | NA | NA | NA | NA | NA | NA | NA | 0.001548 | 0.002183 | <1e-6 | NA | NA | NA | NA | NA | NA | NA | NA | NA | NA | NA | NA | NA | NA | NA | NA | NA | NA | NA | NA | NA |
| PRKN | rs751037529 | chr6:161785793:C:G | NA | NA | NA | NA | NA | NA | NA | NA | NA | NA | NA | NA | NA | NA | NA | NA | NA | NA | 9.68E-05 | <1e-6 | 0.0002033 | NA | NA | NA | NA | NA | NA | NA | NA | NA | NA | NA | NA |
| PRKN | rs377554392 | chr6:161785839:A:T | NA | NA | NA | NA | NA | NA | NA | NA | NA | NA | NA | NA | 0.0005875 | 0.0009524 | <1e-6 | NA | NA | NA | NA | NA | NA | NA | NA | NA | NA | NA | NA | NA | NA | NA | NA | NA | NA |
| PRKN | rs137853059 | chr6:162443314:A:T | NA | NA | NA | NA | NA | NA | 0.0001883 | 0.000387 | <1e-6 | NA | NA | NA | NA | NA | NA | NA | NA | NA | NA | NA | NA | NA | NA | NA | NA | NA | NA | NA | NA | NA | NA | NA | NA |
| PRKN | rs368134308 | chr6:162443356:C:G | NA | NA | NA | NA | NA | NA | NA | NA | NA | NA | NA | NA | NA | NA | NA | NA | NA | NA | NA | NA | NA | 2.58E-05 | 4.72E-05 | <1e-6 | NA | NA | NA | NA | NA | NA | NA | NA | NA |

MAF: minor allele frequency

MAF_A: minor allele frequency of individuals with Parkinson's Disease

MAF_U: minor allele frequency of control individuals
